# Genetic architecture of social anxiety disorder: examining moderation by rural-urban residential environment

**DOI:** 10.64898/2026.11.13.26367930

**Authors:** Regan Harle, Makayla L. Reed, Kyra L. Feuer, Rachel L. Kember

## Abstract

Social anxiety disorder (SAD) is a common psychiatric condition characterized by a consistent fear of social evaluation. Despite heritability estimates of up to 69%, genome-wide association studies (GWAS) of SAD remain limited relative to other psychiatric disorders. Furthermore, the contribution of environmental factors, particularly rural versus urban residence, to SAD risk is poorly characterized. Using the All of Us dataset (N=414,830, Ncases=10,918), our aims were to: (1) identify genetic risk loci for SAD through within-ancestry GWAS (N=1,467-222,500), cross-ancestry meta-analysis, and a European-ancestry meta-analysis (N =788,384); (2) further characterize the genetic architecture of SAD through polygenic risk score (PRS), phenome-wide association (PheWAS), laboratory-wide association (LabWAS), and genetic correlation; and (3) stratify all analyses by rural and urban environment. Across all ancestries examined, rural residence was associated with increased odds of SAD (OR=1.42, 95%; CI [1.36–1.50]; p<5×10). Although no genome-wide significant loci were identified, several loci reached suggestive significance, notably an intronic variant in *NEGR1* (rs12065683), a gene with prior associations to anxiety and neurodevelopmental disorders. PheWAS analyses identified ear disorders as the most nominally significant phenotype (p=3.76×10; OR=1.45) and significant associations with lymphocyte count were identified in the LabWAS (p=1.80×10⁻⁵). These findings provide further insight into the genetic architecture of SAD and suggest that residential environment may modify risk. Larger-scale genetic studies will be required to validate the observed suggestive loci and gene x environment interaction models to form conclusions regarding the true effect of residence.

## Introduction

Social anxiety disorder (SAD), also known as social phobia, is one of the most prevalent and disabling anxiety disorders, affecting approximately 4% of the population worldwide with a lifetime prevalence of 7–12% [1]. Characterized by intense and persistent fear of social situations where the individual may perceive that they are being scrutinized or judged by others, SAD typically emerges during adolescence and, if left untreated, can significantly impair academic, occupational, and social functioning throughout the lifespan [2–5].

Even with heritability estimates reported to be between 13–69% [1], few genetic variants have been identified [6]. More recent, larger-scale efforts focused on a group of anxiety disorders, referred to as major anxiety disorders (ANX), have substantially advanced the field: a genome-wide association study (GWAS) of ANX in 122,341 European-ancestry cases identified 58 genome-wide significant loci and implicated GABAergic signaling as a central biological pathway [7], while a complementary meta-analysis of quantitatively measured general anxiety disorder (GAD) symptoms further delineated the polygenic architecture of anxiety-related traits across European-ancestry cohorts [8]. Other analyses, including those leveraging the UK Biobank (UKBB) [9], the Million Veteran Program (MVP) [10], and the Psychiatric Genetics Consortium (PGC) [11], have established substantial polygenicity for anxiety-related traits and significant genetic correlation across major anxiety subtypes and with major depressive disorder (MDD), consistent with a shared internalizing liability spectrum. In addition, polygenic risk scores (PRS) derived from ANX GWAS confer modest but significant prediction of anxiety-related outcomes [9], yet whether this polygenic signal could also reflect SAD-specific liability has not been evaluated. SAD has been shown to be phenomenologically distinct from GAD by its stimulus specificity rather than the pervasive, difficult-to-control worry across multiple life domains that defines the latter [2], and the near-exclusive focus on European-ancestry samples limits locus discovery and likely underestimates the genetic diversity underlying SAD risk [11, 12].

The environment can also be a source of risk for SAD [9,13]. The geographic environment, namely urban vs. rural residency, has been proven as a potential factor of psychiatric risk in younger people. Specifically, rural residence may contribute to greater odds of having SAD in adolescents [14,15], with this being extended into adults through National Health surveys [16]. However, some studies in adults have instead shown an increased risk of social stress with urban residence [17–19]. This coincides with the urban stress hypothesis [20], which proposes that urban residency could be associated with elevated rates of schizophrenia, depression and overall stress due to social density, economic inequality, noise, and social fragmentation [21,22]. For SAD, however, the direction of any rurality effect is theoretically equivocal, as, for instance, sparse social environments may restrict habituation opportunities and reinforce avoidance behaviors central to cognitive-behavioral models of SAD maintenance [2]. This social facilitation model proposes that regular exposure to social situations provides opportunities for habituation and development, potentially protecting against the development of social anxiety [23]. Alternately, rural communities often feature tighter social networks where social evaluation may be more salient and persistent, potentially exacerbating social anxiety symptoms among vulnerable individuals [24]. These competing hypotheses have not been rigorously evaluated in large, ancestrally diverse cohorts. In fact, the disorder appears to manifest across diverse cultural contexts, though its expression and prevalence may vary based on sociocultural factors and environmental conditions [25]. Furthermore, geographic environment may modulate genetic risk, and, therefore, play a moderating role in the development of SAD.

To understand the complex etiology of SAD, we conducted a within- and cross-ancestry GWAS of SAD using the All of Us (AoU) Research Program [26], followed by within-European ancestry meta-analysis incorporating summary statistics from the UKBB [27] and FinnGen [28], to uncover loci associated with SAD. AoU GWAS are further stratified by rural versus urban residence to test whether geographic context modifies SAD genetic risk. We additionally perform PRS analyses, sex-stratified phenome-wide association studies (PheWAS), lab-wide association studies (LabWAS), and genetic correlation analyses to compare the genetic and phenotypic architecture of SAD to that of ANX.

## Subjects and Methods

### Study Population and Phenotype Definitions

Participants came from the AoU dataset (controlled tier, v8, accessed June 2025). The AoU Research Program is a longitudinal cohort study that has enrolled a diverse group of participants across the United States [26]. We utilized the Controlled Tier dataset, which provides access to clinical-grade genome sequences and comprehensive phenotypic data [26].

Two case definitions for SAD were applied. The clinical definition required at least one International Classification of Diseases, Tenth Revision (ICD-10) code for social phobia (F40.10 or F40.11) in the participant’s electronic health record (EHR). The survey-based definition required affirmative responses to all three SAD-relevant items in the AoU Behavioral Health and Personality survey, derived from Diagnostic and Statistical Manual of Mental Disorders, 5th Edition (DSM-5) diagnostic criteria: (i) *“Were you ever in your life so painfully shy or scared of social situations (such as attending a party, speaking to strangers, or eating in public) that you avoided social situations whenever you could?”*; (ii) *“Do you worry more than most people about being embarrassed, looking stupid, or being judged by other people?”*; and (iii) *“Did this ever last for more than 6 months?”.* Individuals were considered cases if they met either the clinical or survey-based definition. Controls were defined as individuals with no ICD-10 codes for social phobia and no affirmative responses to any of the three survey items. All analyses were conducted independently within six genetically inferred ancestral groups: African (AFR), Admixed American (AMR), East Asian (EAS), European (EUR), Middle Eastern (MID), and South Asian (SAS).

### Genomic Data and Quality Control

Quality control (QC) of the AoU dataset has been previously described [29] and participants were assigned to genetic ancestry groups by projection onto the 1000 Genomes Project Phase 3 reference panel [30] via PCA, integrated with self-reported ancestry in AoU. In addition to this, we excluded flagged variants, which were noted as those failing the QC most likely due to poorly-sampled ancestry, relatedness pruning (removing all individuals listed in the AoU provided kinship file), excluded variants with MAF<1%, excluded variants violating HWE (p<1×10^-6^) in controls, performed ancestry-specific principal component analysis (PCA) and included biallelic SNPs only in the analysis. Variant- and sample-level QC was implemented using Hail v0.2 (Broad Institute, Cambridge, MA; https://github.com/broadinstitute/hail) variant_qc and sample_qc functions.

### Genome-Wide Association Study

All analyses were implemented in Hail v0.2 and PLINK 2.0 [31]. Within each ancestral group, logistic regression under an additive genetic model was used to test the association between each single-nucleotide polymorphism (SNP) and SAD case/control status, adjusting for age, sex, and the first 10 within-ancestry principal components (PCs). Genome-wide significance was defined as p<5×10□□; a suggestive threshold of p<1×10□□ was additionally applied. Genomic inflation was assessed using the inflation factor λ.

### Meta-analysis

Cross-ancestry meta-analysis was performed by combining ancestry-stratified GWAS summary statistics using inverse-variance weighted (IVW) fixed-effects meta-analysis implemented in METAL [32]. Heterogeneity across ancestry-stratified results was quantified using Cochran’s Q statistic and I².

To maximize statistical power within a genetically homogeneous context, a EUR meta-analysis using METAL was additionally conducted combining AoU EUR summary statistics with SAD GWAS summary statistics from the UKBB [27] and FinnGen [28].

### Geographic Classification

Participant residential ZIP3 codes were linked to Rural-Urban Commuting Area (RUCA) codes (available at https://www.ers.usda.gov/data-products/rural-urban-commuting-area-codes/), a US Department of Agriculture (USDA) classification system that categorizes census tracts based on population density, urbanization, and daily commuting patterns [33,34]. For primary analyses, a binary scheme distinguished urban (RUCA codes 1–3) from rural (RUCA codes 4–10) residence, consistent with established conventions in previous research [35–40].

### Socioeconomic Variables

Socioeconomic data were extracted from the AoU Socioeconomic Status tables. Measures included the Area Deprivation Index (ADI), a validated composite of neighborhood-level disadvantage derived from US Census data, encompassing income, educational attainment, employment, and housing quality [41] and ZIP-code-level median household income.

### Rural-Urban Association Analyses

Ancestry-stratified logistic regression was used to estimate associations between rural residence and SAD risk. Within each ancestry group, a multivariable logistic regression model was fitted using maximum likelihood estimation. The binary outcome was SAD status. Four models were tested: (1) unadjusted; (2) adjusted for age and sex; (3) additionally adjusted for the first 10 within-ancestry PCs; and (4) adjusted for ADI and median household income. The regression coefficient corresponded to rural residence. Ancestry-specific effect estimates were combined using IVW fixed-effect meta-analysis.

### Polygenic Risk Score Construction

PRS were constructed using EUR meta-analysis SAD GWAS summary statistics and ANX GWAS as the discovery sample. SNP weights were estimated using PRS-CS [42] and the 1000 Genomes Project EUR reference panel. The target cohort was the Penn Medicine Biobank (PMBB) EUR ancestry individuals. Target sample genotype data were subjected to standard QC procedures as described previously [43]. PRS were standardized prior to analysis.

### Phenome-Wide Association Study

To characterize the phenotypic consequences of genetic liability for SAD, a PheWAS was conducted in the PMBB using the standardized SAD and ANX PRS. Clinical phenotypes were defined by mapping ICD-9 and ICD-10 diagnosis codes to PheCodes (version 1.2) using established mapping and exclusion criteria [44]. Logistic regression was performed under anadditive model, adjusting for age, sex, and the first 10 genetic PCs, with sex-stratified analyses conducted in parallel. A Bonferroni-corrected significance threshold (p<0.05/1816 phenotypes tested was applied; nominal associations (p<0.05) are additionally reported. Analyses were implemented using the PheWAS R package (version 0.99.6) [45].

### Laboratory-Wide Association Study

A LabWAS was performed to evaluate associations between genetic liability for SAD and ANX and quantitative clinical laboratory measures extracted from the PMBB. Linear regression was conducted with each laboratory value as the outcome and the standardized SAD and ANX PRS as the primary predictor, adjusting for age, sex, and the first 10 genetic PCs; sex-stratified analyses were performed in parallel. To account for multiple testing across laboratory traits, statistical significance was defined using a Bonferroni threshold of 0.05/79 tests. Nominal associations (p*<*0.05) were also reported for exploratory purposes.

### Genetic Correlation Analysis

Genetic correlations between SAD and ANX and a broad range of external phenotypes were estimated using LD score regression (LDSC) [46] via the Complex Trait Genetics Virtual Lab (CTG-VL); accessed August 2026; https://vl.genoma.io/) [47]. SAD summary statistics were drawn from the EUR meta-analysis; comparison trait summary statistics were obtained from the traits catalogued in CTG-VL. For each pairwise comparison, the genetic correlation coefficient (rg), standard error, and p-value were estimated. Statistical significance was defined using a Bonferroni correction for the number of traits tested (p<0.05/3196 traits); nominal associations are additionally reported to facilitate interpretation of broader genetic correlation patterns.

## Results

### Cohort Demographics

GWAS were conducted within ancestral group in the AoU dataset for 74,576 AFR-like ancestry individuals (cases=600, controls=73,976), 70,626 AMR-like ancestry individuals (cases=908, controls=69,718), 9707 EAS-like ancestry individuals (cases=181, controls=9526), 222,500 EUR-like ancestry individuals (cases=8497, controls=214,003), 1467 MID-like ancestry individuals (cases=29, controls=1438) and 5368 SAS-like ancestry individuals (cases=104, controls=5264). Cases were significantly younger, more likely to be female and have a lower ADI, with no differences between cases and controls in income (Table 1).

**Table 1.**
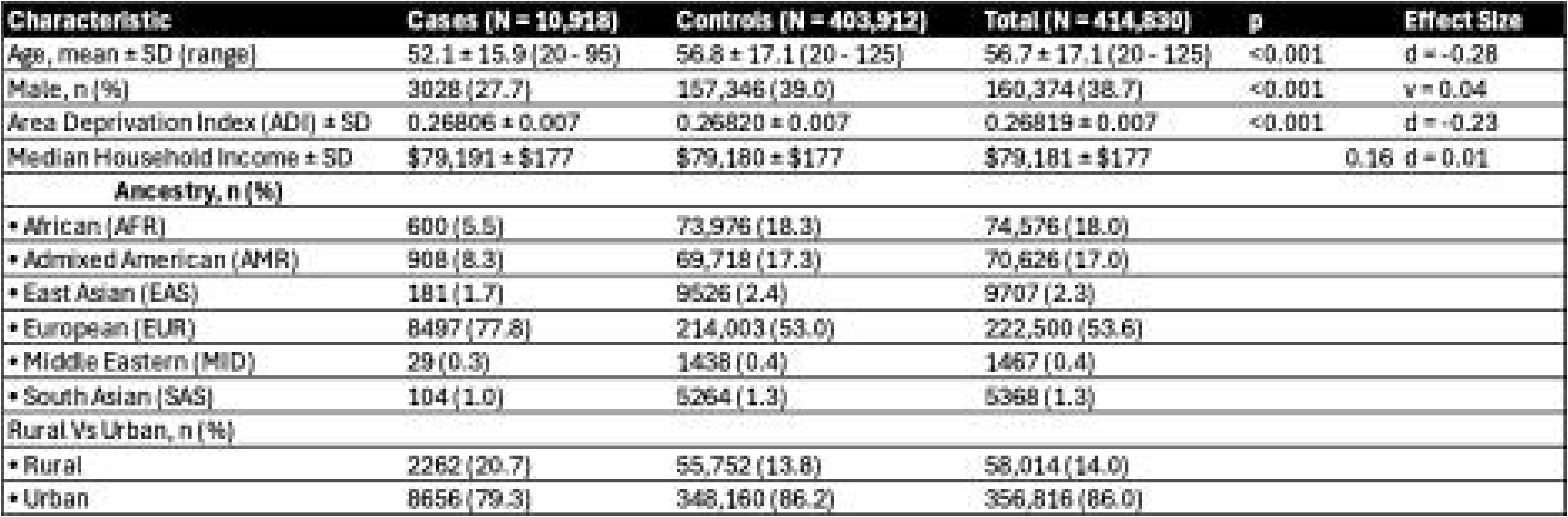
Demographics of the SAD sample used for the analyses from the AoU V8 dataset.

| Characteristic | Cases (N = 10,918) | Controls (N = 403,912) | Total (N = 414,830) | p | Effect Size |
|---|---|---|---|---|---|
| Age, mean ± SD (range) | 52.1 ± 15.9 (20 - 95) | 56.8 ± 17.1 (20 - 125) | 56.7 ± 17.1 (20 - 125) | <0.001 | d = -0.28 |
| Male, n (%) | 3028 (27.7) | 157,346 (39.0) | 160,374 (38.7) | <0.001 | v = 0.04 |
| Area Deprivation Index (ADI) ± SD | 0.26806 ± 0.007 | 0.26820 ± 0.007 | 0.26819 ± 0.007 | <0.001 | d = -0.23 |
| Median Household Income ± SD | $79,191 ± $177 | $79,180 ± $177 | $79,181 ± $177 | 0.16 | d = 0.01 |
| **Ancestry, n (%)** |  |  |  |  |  |
| ▪ African (AFR) | 600 (5.5) | 73,976 (18.3) | 74,576 (18.0) |  |  |
| ▪ Admixed American (AMR) | 908 (8.3) | 69,718 (17.3) | 70,626 (17.0) |  |  |
| ▪ East Asian (EAS) | 181 (1.7) | 9526 (2.4) | 9707 (2.3) |  |  |
| ▪ European (EUR) | 8497 (77.8) | 214,003 (53.0) | 222,500 (53.6) |  |  |
| ▪ Middle Eastern (MID) | 29 (0.3) | 1438 (0.4) | 1467 (0.4) |  |  |
| ▪ South Asian (SAS) | 104 (1.0) | 5264 (1.3) | 5368 (1.3) |  |  |
| Rural Vs Urban, n (%) |  |  |  |  |  |
| ▪ Rural | 2262 (20.7) | 55,752 (13.8) | 58,014 (14.0) |  |  |
| ▪ Urban | 8656 (79.3) | 348,160 (86.2) | 356,816 (86.0) |  |  |

### Within-Ancestry Genome-Wide Association Study

Within-ancestry GWAS identified 19 loci at suggestive significance in AFR, 17 loci in AMR, 15 loci in EAS, 10 in EUR, and 2 loci in SAS (Supplementary Tables 1-5, Supplementary Figures 1-5). Among these, the most significantly shared loci between ancestries were those in the region of chromosome 2 in AMR and EUR ancestries (lead SNP:rs12468520 in AMR and rs11897777 in EUR), and those in the chromosome 4 locus region in EAS and EUR ancestries (lead SNP:rs223976 in AMR and rs3931694 in EUR).

### Meta-Analyses

#### Cross-ancestry

The cross-ancestry meta-analysis consisted of 414,830 individuals (cases=10,918, controls=403,912) and identified 18 LD-independent loci at suggestive significance (p<1×10□□; Figure 1; Supplementary Table 6). The strongest association was an exonic variant in *PIKFYVE*(rs999891; p=1.52×10□□), nominally significant in both AMR and EUR ancestries, with consistent direction of effect across ancestry groups. Additional suggestive associations included an intronic variant in *NEGR1* (rs12065683; p=7.54×10□□).

**Figure 1.**
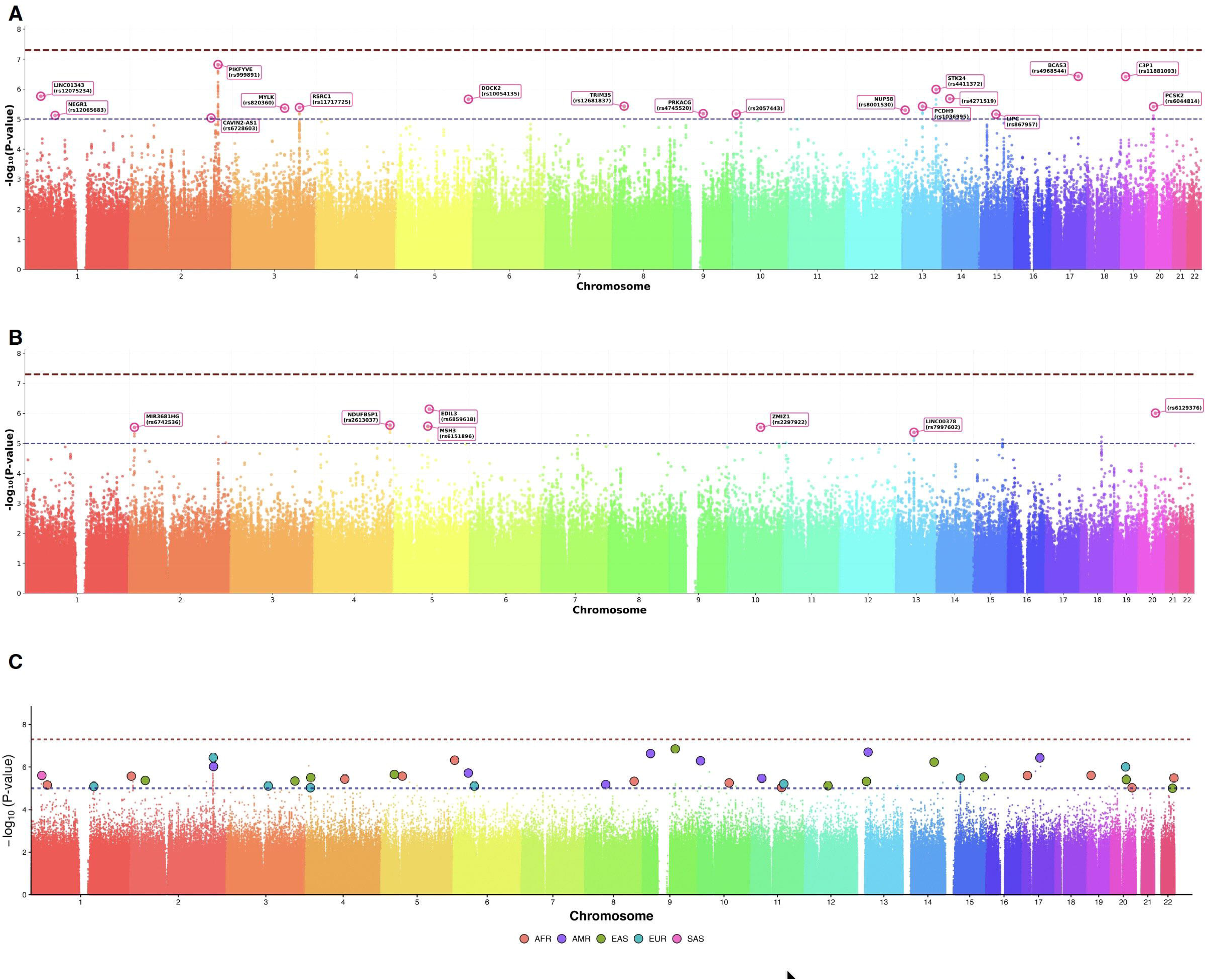
Manhattan plots showing genome-wide association results for the social anxiety phenotype in the All of Us cohort (ICD10 and/or answering ‘yes’ on three social anxiety survey questions). The y-axis represents −log₁₀(p-values), and the x-axis represents chromosomal positions. The dark red dashed line denotes genome-wide significance (p<5×10⁻⁸); the blue dashed line denotes suggestive significance (p<1×10⁻⁵). Top loci are annotated with nearest gene symbols. (a) meta-analyzed across ancestries (b) EUR ancestry meta-analysis (including AoU, FinnGen and UKB) (c) porcupine plot showing the top LD clumped SNPs per ancestry.

#### EUR Only

The EUR meta-analysis comprised 788,384 individuals (cases=14,209, controls=774,175) across AoU (N=222,500), FinnGen (N=448,168), and UKBB (N=117,716). Seven LD-independent loci reached suggestive significance (p<1×10□□), with the strongest association observed with an intronic variant in *EDIL3* (rs6859618; p=7.4×10□□), followed by an intergenic variant near *NDUFB5P1* (rs2613037; p=2.5×10□□), and intronic variants in *MSH3* (rs6151896; p=2.7×10□□); and *LINC00378* (rs7997602; p=4.3×10□□; Figure 1; Supplementary Table 7).

### Polygenic Risk Score

PRS for SAD (SAD-PRS) and major anxiety disorders (ANX-PRS) were tested for association with ANX and SAD (Supplementary Figure 7). SAD-PRS did not reach statistical significance for either outcome, showing only suggestive association with social anxiety, although in the expected direction (OR=1.020, 95% CI: 0.999–1.042, p=0.067) and no association with major anxiety disorders (OR=1.014, 95% CI: 0.992–1.036, p=0.261). ANX-PRS significantly predicted both ANX (OR=1.118, 95% CI: 1.094–1.141, p<0.0001) and SAD (SAD; OR=1.081, 95% CI: 1.058–1.105, p=0.001).

### Phenome-Wide Association Study

Although no associations survived Bonferroni correction for SAD-PRS, the strongest associations were with disorders of the middle ear and mastoid (OR=1.45, p=3.76×10□□) and sleep apnea (OR=1.06, p=6.79×10□□), alongside associations across the mental health domain including mood disorders, MDD, and substance use disorders (SUDs) (Figure 2a; Supplementary Table 8). Sex-stratified analyses revealed that the ear/mastoid and sleep apnea associations were predominantly driven by males (OR=1.57, p=2.70×10□□ and OR=1.07, p=3.72×10□□, respectively), whereas in females the top associations were with complications of trauma or procedure (OR=0.76, p=9.70×10□□) and chronic pain syndrome (OR=0.84, p=0.001) (Supplementary Figure 8-9; Supplementary Table 9-10).

**Figure 2.**
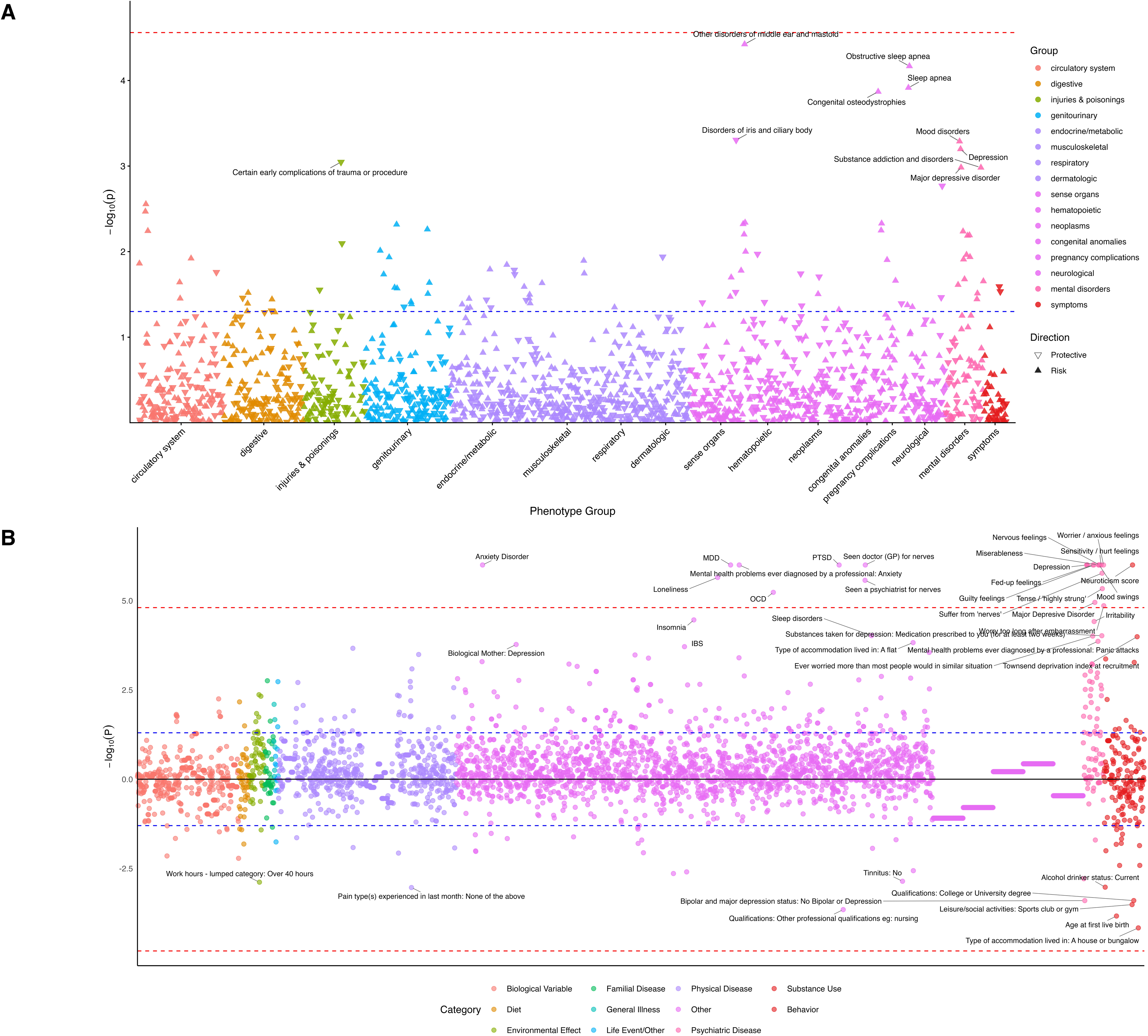
(a) Manhattan plot of the PheWAS results against SAD, with each point representing an individual phenotype, grouped according to the corresponding Phecode 1.2 disease system (with a minimum of two phecodes and exclusion map true) and colored by phenotype category. The blue dashed line denotes Bonferroni significance (p<0.05/1816 phenotypes tested); the dark red dashed line denotes nominal significance (p<1×10). Upward-pointing triangles indicate risk associations, whereas downward-pointing triangles indicate protective associations. (b) Genetic correlation analysis on different traits compared to SAD, generated by the Complex Traits Genetics Virtual Lab (CTG-VL) in European ancestry. The y-axis represents the log-transformed false discovery rate–corrected p values across the traits included in the analysis, with the dashed line representing the significance threshold.

In contrast to the SAD PheWAS, the PheWAS for ANX-PRS showed numerous significant associations, with the strongest signals observed within the mental health domain (Supplementary Table 11). As expected, the most significant associations were with anxiety-related phenotypes, including ANX (OR =1.19, p=2.18×10□□) and GAD (OR=1.20, p=1.68×10□¹□). Strong associations were also observed with several psychiatric comorbidities, including mood disorders (OR= 1.19, p=1.54×10□³□), depression (OR=1.20, p=1.75×10□³□), and bipolar disorder (OR=1.23, p=9.83×10□¹□). Beyond psychiatric phenotypes, the ANX-PRS demonstrated widespread associations across multiple organ systems. Significant associations were observed with gastrointestinal disorders, including gastroesophageal reflux disease (GERD); OR=1.11, p=7.14×10□¹□) and related oesophageal diseases (OR=1.10, p=1.17×10□¹□), respiratory conditions such as chronic airway obstruction (OR=1.15, p=5.02×10□¹□), asthma (OR =1.12, p=3.18×10□¹□) and chronic bronchitis (OR=1.18, p=8.88×10□¹□).

### Genetic Correlations

Genetic correlation analyses identified 22 Bonferroni-significant associations of SAD with external traits (Figure 2b; Supplementary Table 12). The strongest positive correlations were with all anxiety disorders (rg=0.666, SE=0.113, p<0.001), neuroticism (rg=0.631, SE=0.104, p<0.001), and sensitivity/hurt feelings (rg=0.619, SE=0.111, p<0.001). Substantial correlations were also observed with post-traumatic stress disorder (PTSD); rg=0.515, SE=0.094, p<0.001), depression (rg=0.524, SE=0.097, p<0.001), and MDD (rg=0.512, SE=0.093, p<0.001).

Comparatively, a genetic correlation analysis of ANX identified >100 Bonferroni-significant associations with external traits (Supplementary Figure 10; Supplementary Table 13). Among these, the strongest positive correlations were, similar to the SAD findings, neuroticism (rg=0.695, SE=0.03, p<0.001) and seeing a psychiatrist for ‘nerves’ or ‘worry’ (rg=0.845, SE =0.03, p<0.001). But, differently to SAD, ANX was most highly positively correlated with mood swings (rg=0.651, SE=0.03, p<0.001), dorsalgia (rg=0.496, SE=0.02, p<0.001) and GERD (rg=0.512, SE=0.02, p<0.001).

A forest plot comparison of SAD and ANX’s genetic correlations of some of the most significant traits is shown in Supplementary Figure 11.

### Laboratory-Wide Association Study

LabWAS across 79 laboratory traits for SAD-PRS identified significant positive associations with lymphocyte count (N=30,686; β=0.02, SE=0.006, p=1.80×10⁻⁵) and significant negative associations with height (N=35,011; β=−0.02, SE=0.004, p=4.41×10⁻⁴), and a nominally significant positive association with white blood cell count (WBC) (N=33,594; β=0.02, SE=0.006, p=0.001), (Figure 3, Supplementary Table 14). Sex-stratified analyses revealed that the lymphocyte count, WBC, and inverse height associations were present in both sexes; males additionally showed a positive association with body mass index (BMI) (N=17,950; β=0.02, SE=0.006, p=0.003) and females with body temperature (N=15,188; β=0.02, SE=0.008, p=0.02) (Supplementary Figure 12, Supplementary Tables 15-16).

**Figure 3.**
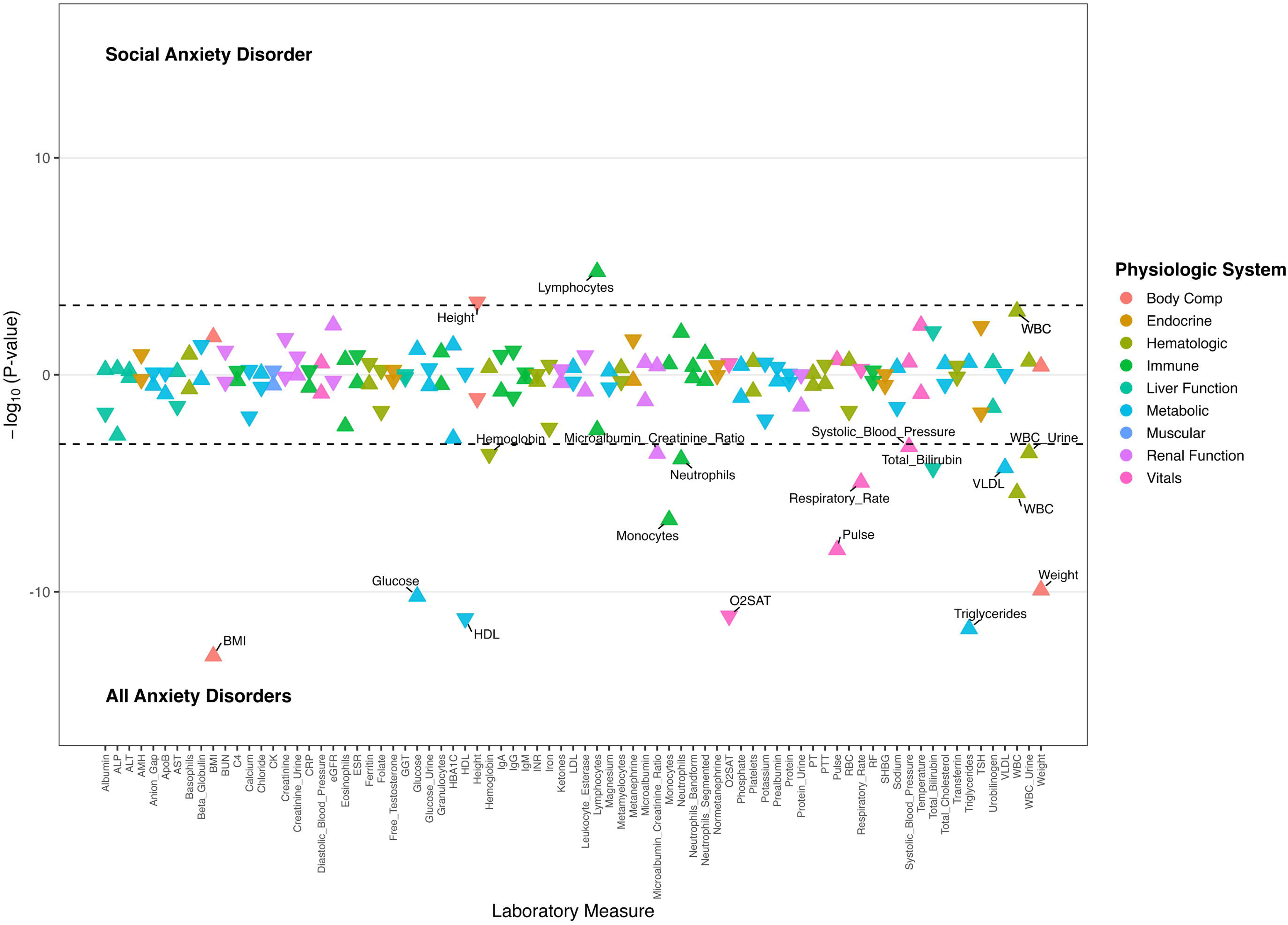
Comparative laboratory-wide association study (LabWAS) results for social anxiety disorder (SAD) and major anxiety disorders (ANX) in individuals of European ancestry. Laboratory associations are displayed as a mirrored Hudson-style plot, with SAD shown above the horizontal axis and ANX shown below the horizontal axis. Each laboratory measure is represented by a directional triangle, where ▴ indicates a positive effect estimate (higher laboratory values associated with greater disorder risk) and ▾ indicates a negative effect estimate (lower laboratory values associated with greater disorder risk). Colors denote laboratory system categories. Dashed horizontal lines indicate Bonferroni-corrected significance thresholds for each analysis (p<0.05/79 tests). Laboratory measures meeting Bonferroni significance are labeled by name. WBC indicates white blood cell count; eGFR estimated Glomerular Filtration Rate; HBA1C Hemoglobin A1c; TSH Thyroid-Stimulating Hormone.

LabWAS for ANX-PRS showed the strongest associations for anthropometric and metabolic traits, including a positive association with BMI (N=34,045, β=0.04, SE=0.005, p=1.09×10 ¹³), triglycerides (N=21,277, β=0.05, SE=0.007, p=1.95×10 ¹²), and glucose levels (N=33,829, β =0.04, SE=0.005, p=6.23×10 ¹¹) and a negative association with lower high-density lipoprotein (HDL) cholesterol (N=20,581, β=-0.05, SE=0.007, p=5.87×10 ¹²) (Supplementary Figure 13; Supplementary Table 17).

### Rural-Urban Stratification

#### Rural Residence and SAD Risk

Logistic regression analyses across the full AoU sample revealed a robust, highly significant positive association between rural residence and SAD risk that persisted across all four models (Table 2).

**Table 2.**
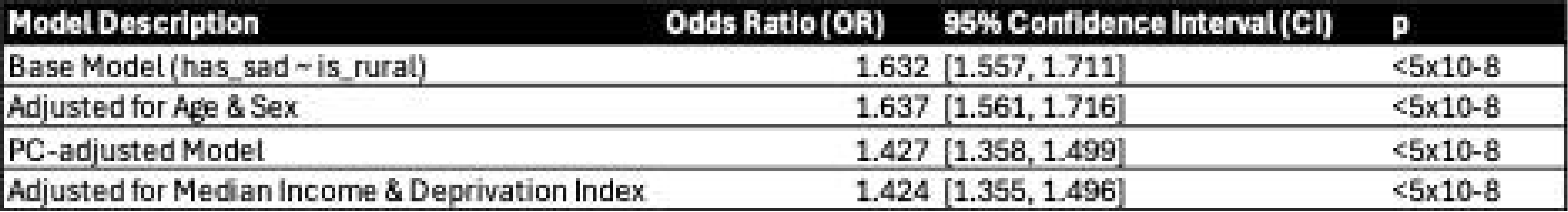
Comparison of Logistic Regression models for rural effect on social anxiety.

| **Model Description** | **Odds Ratio (OR)** | **95% Confidence Interval (CI)** | **p** |
|---|---|---|---|
| Base Model (has_sad ~ is_rural) | 1.632 | [1.557, 1.711] | <5x10-8 |
| Adjusted for Age & Sex | 1.637 | [1.561, 1.716] | <5x10-8 |
| PC-adjusted Model | 1.427 | [1.358, 1.499] | <5x10-8 |
| Adjusted for Median Income & Deprivation Index | 1.424 | [1.355, 1.496] | <5x10-8 |

In the unadjusted model, rural residence was associated with 63% greater odds of SAD (OR = 1.632, 95% CI [1.557–1.711], p<5×10□□). This association was essentially unchanged after adjustment for age and sex (OR=1.637, 95% CI [1.561–1.716], p<5×10□□). Adjustment for the first 10 within-ancestry PCs attenuated the effect modestly (OR=1.427, 95% CI [1.358–1.499], p<5×10□□), suggesting a partial contribution of population stratification. However, the association remained highly significant and of substantial magnitude. Further adjustment for ADI and median household income produced negligible additional attenuation (OR=1.424, 95% CI [1.355–1.496], p<5×10□□), indicating that the rural–SAD association is not primarily mediated by socioeconomic disadvantage as captured by these indicators. Ancestry-stratified ORs are shown in Supplementary Figure 14. While confidence intervals were wider in smaller ancestry groups, the direction of effect was consistent across all groups.

#### Rural and Urban Stratified GWAS

GWAS stratified by rural and urban residence were conducted to examine whether the genetic architecture of SAD differs according to geographic context (Figure 4, Supplementary Tables 18-19).

**Figure 4.**
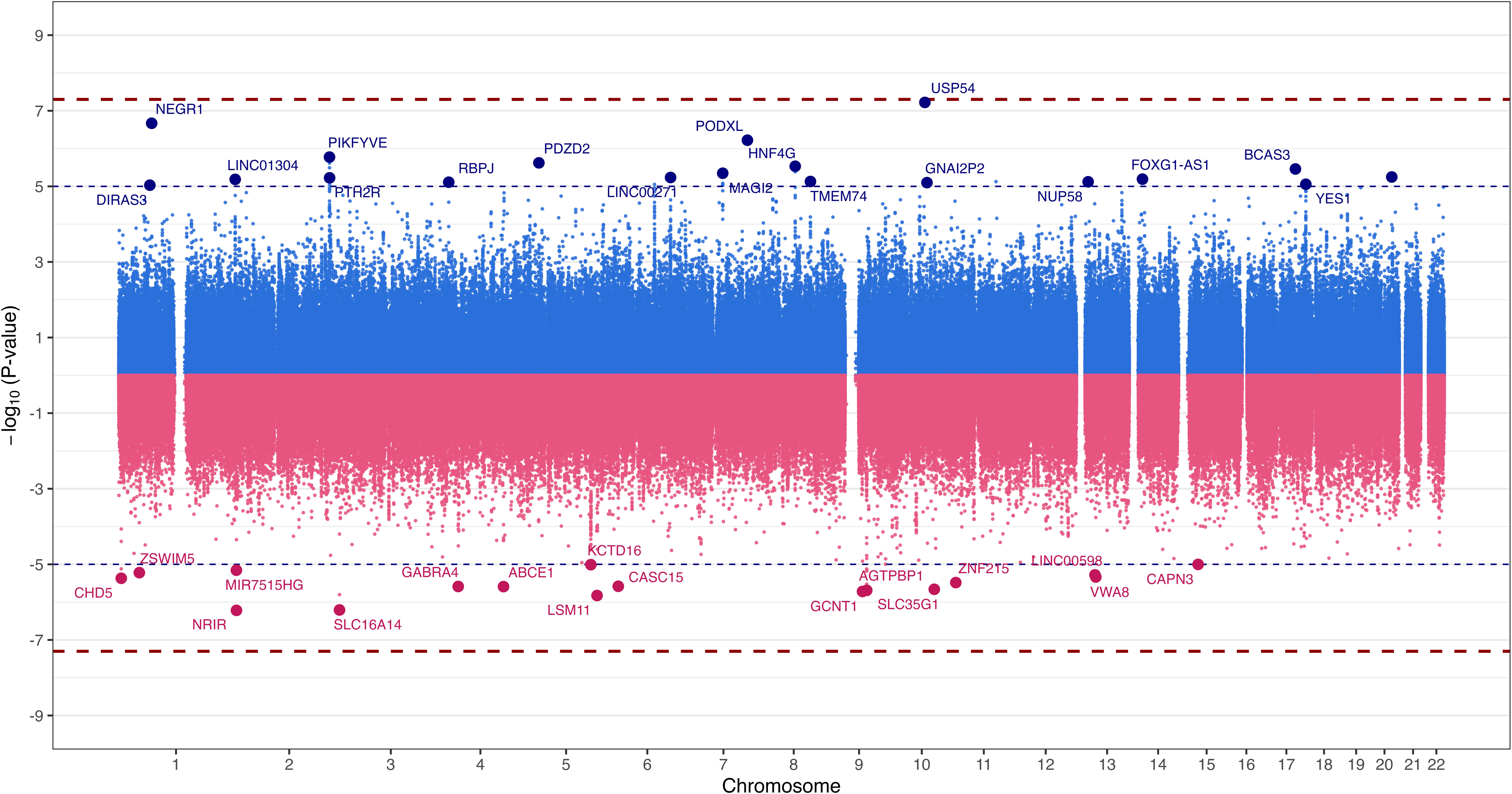
Hudson plot comparing genome-wide association study (GWAS) results from urban and rural ancestry-stratified analyses. Each point represents a single genetic variant plotted according to chromosomal position on the x-axis and association significance on the y-axis as ±log_10_(P). Urban GWAS results are displayed above the x-axis in blue, whereas rural GWAS results are mirrored below the x-axis in pink. Chromosomes 1-22 are shown sequentially along the genome. The orange dotted lines indicate the suggestive significance threshold (p<1×10), and the red dashed lines indicate the conventional genome-wide significance threshold (p<5×10). Labeled loci correspond to independent lead variants identified through linkage disequilibrium (LD)-based clumping and meeting the suggestive significance threshold. For loci mapping to the same nearest gene, only the most significant clumped variant was annotated. Gene labels indicate the nearest annotated gene to each lead variant. No variants reached genome-wide significance in either ancestry-stratified analysis.

In the rural-stratified analysis, 17 independent LD-clumped loci surpassed the suggestive significance threshold. The strongest association was observed near *NRIR* (rs10495544; p=6.04×10□□), followed by variants near *SLC16A14* (rs13407773; p=6.20×10□□) and *LSM11* (rs10056356; p=1.49×10□□).

GWAS for SAD among urban residents identified 19 independent lead loci exceeding the suggestive significance threshold. The strongest association signal was observed near *MAGI2* (rs6466036; p=4.49×10^-6^). Additional lead loci were identified near *PDZD2* (rs7725382; p=2.39×10^-6^).

#### Rural and Urban Heterogeneity Analyses

Between-group heterogeneity was conducted in rural versus urban residence. Several loci demonstrated exceptionally strong evidence of heterogeneity, with all top signals exhibiting I² values exceeding 94% and highly significant heterogeneity tests (p<1×10□□), indicating some differences in effect estimates between rural and urban populations (Supplementary Table 20).

The strongest heterogeneous signal mapped to *CASC15* (rs2328715) which demonstrated an I² of 95.6% (Q=22.566, p=2.03×10□□). The direction of effect was discordant between groups (−/+), suggesting that the effect differed according to residential environment. Likewise, another notable heterogenous signal localized to *LMX1B* (rs7874916) (I²=94.9%, Q=19.699, p=9.06×10□□), again, showing a difference in directionality.

#### Rural and Urban Stratified PheWAS and LabWAS

A PheWAS was also conducted separately on the rural and urban results, however, there were no statistically significant findings (Supplementary Tables 21-22). The LabWAS found different suggestively significant results between rural and urban residence stratified SAD-PRS (Supplementary Tables 23-24). For instance, the most significant finding in the positive direction in the rural setting was granulocyte count (N=3408; β=0.04, SE=0.02, p=0.02) followed by weight (N=35,111; β=0.01, SE=0.005, p=0.03) and in the negative direction albumin count (N=29,902; β =-0.01, SE=0.006, p=0.02) (Supplementary Figure 15), whereas in the urban setting the most significant result in the positive direction was Aspartate Aminotransferase (AST) count (N=30,059; β=0.01, SE=0.006, p=0.01) with creatinine urine being the most significant in the negative direction (N=9202; β=-0.02, SE=0.01, p=0.04) (Supplementary Figure 16).

#### Rural and Urban Stratified Genetic Correlation

Genetic correlation analyses were unable to be conducted in the rural stratified analysis due to the small sample size. However, urban stratified genetic correlation analyses identified 8 Bonferroni-significant associations with external traits (Supplementary Figure 17; Supplementary Table 25). As expected, the majority of positive correlations were with psychiatric-related traits such as depression (rg=0.245, SE=0.05, p<0.001), MDD (rg =0.251, SE=0.05, p<0.001) and ‘worry too long after embarrassment’ (rg=0.318, SE =0.07, p<0.001). The strongest negative correlations were with the ‘age of first episode of depression’ (rg= −0.811, SE=0.16, p<0.001), going to a ‘pub or club’ for social leisure activities (rg=-0.390, SE=0.08, p<0.001) and ‘lack of illnesses in siblings’ (rg=-0.319, SE=0.07, p<0.001).

## Discussion

This work provides a comprehensive genetic characterization of SAD as a distinct phenotype within the anxiety spectrum, integrating cross-ancestry GWAS, European-ancestry meta-analysis, PheWAS, LabWAS, and genetic correlation analyses in one of the largest and most ancestrally diverse SAD datasets assembled to date.

Cross-ancestry meta-analyses and the urban stratified GWAS both identified a nominally significant locus containing *NEGR1*. *NEGR1* is a member of the IgLON family of neuronal cell adhesion molecules, regulates synaptic formation and neuronal connectivity, and has been implicated in MDD [48], ASD [49,50], and cognitive ability [51,52] Additionally, one of the largest meta-analyses to date of ANX found genome-wide significant loci containing *NEGR1* [7] and in various mouse studies of the brain *negr1* has also been shown to play a role in anxiety and depression phenotypes [53,54].

The most significant finding from the cross-ancestry meta-analysis, loci containing *PIKFYVE,* was also found in EUR and AMR. *PIKFYVE* is involved in endosomal-lysosomal trafficking and this could possibly be a contributor to the increased lymphocyte count found in people with SAD [55]. In the EUR meta-analysis, the top associated loci contained *EDIL3*, which encodes an integrin-binding extracellular matrix protein, and has been shown to be an important regulator of synaptic plasticity and neural circuit remodeling [56].

PheWAS analysis revealed associations between SAD PRS and a range of phenotypes spanning both psychiatric and physical, although none passed multiple testing correction. The most significant association was with disorders of the middle ear, which could be noteworthy from a developmental standpoint. Acquired middle ear disease frequently presents or is first diagnosed during adolescence [57], the same developmental window in which SAD typically emerges [1,2]. PheWAS also found an association with sleep apnea, which is consistent with the clinical overlap between anxiety disorders and sleep-disordered breathing, mediated in part by shared autonomic dysregulation [58]. To our knowledge, this association has not been previously reported at the polygenic level for SAD specifically. Co-occurrence with MDD and SUDs recapitulates established clinical comorbidity patterns [60,61] and is consistent with the shared internalizing genetic architecture. The sex-stratified results, with ear/mastoid and sleep apnea associations concentrated in males and pain-related phenotypes prominent in females, are in keeping with documented sex differences in the somatic expression of anxiety [61] and suggest that sex-differentiated biological pathways may mediate distinct phenotypic consequences of SAD genetic liability.

In comparison, ANX PRS demonstrated robust associations across disorders that are well established as part of the broader anxiety comorbidity spectrum, including depression, PTSD, bipolar disorder, TUD, GERD and asthma [53,63–65]. The most significant association in the SAD-PRS, disorders of the middle ear, was not significant in the ANX PRS.

LabWAS found elevated lymphocyte and WBC counts associated with SAD polygenic risk, which adds to an emerging body of evidence linking anxiety disorders to systemic immune dysregulation. Chronic social-evaluative stress, a defining feature of SAD, activates both the Hypothalamic-Pituitary-Adrenal (HPA) axis and sympathoadrenal pathways, with downstream effects on lymphocyte trafficking and proliferation [65–67]. Critically, these associations reflect constitutional genetic propensity rather than acute psychological state, suggesting that neuroimmune perturbation may represent a stable biological substrate of social anxiety liability rather than a purely syndromal epiphenomenon. The inverse polygenic association with height, present in both sexes, is consistent with prior cross-trait analyses identifying negative genetic correlations between internalizing phenotypes and anthropometric traits, while the male-specific positive association with BMI parallels epidemiological observations of sex-differentiated metabolic correlates of anxiety.

The LabWAS overlap between SAD and ANX was modest. Elevated WBC count was one of the few findings shared across both ANX and SAD, suggesting that immune-related mechanisms may represent a common biological feature across anxiety phenotypes. This interpretation is consistent with accumulating evidence linking anxiety symptoms and anxiety disorders to inflammatory and immune dysregulation, including elevated peripheral inflammatory markers and alterations in circulating immune cell population [69,70]. In addition, the ANX-PRS was characterized by multiple markers of metabolic dysfunction. These findings align with evidence from epidemiological and meta-analytic studies demonstrating links between anxiety disorders and metabolic syndrome, obesity, dyslipidemia, and impaired glucose regulation [63]. Recent longitudinal work has further shown that elevated triglycerides, glucose and lower HDL levels are associated with increased future risk of anxiety and related stress disorders, suggesting that metabolic dysregulation and anxiety may share common biological pathways [70]. Since these are not significant within SAD alone, it could suggest that different biological mechanisms are at play in SAD [72,73].

Genetic correlation analyses confirm that SAD lies within the internalizing spectrum while retaining a degree of genetic individuality. The high positive correlations with neuroticism and general anxiety are expected given the extensive genetic sharing across this domain [53]. Neuroticism, particularly, has emerged as a key transdiagnostic genetic liability factor for both anxiety and depressive disorders [73]. The significant correlations with PTSD, MDD, and depression align with the established comorbidity structure of internalizing conditions [60] and are consistent with dimensional models of psychopathology positing a general internalizing factor [74]. Notably, the genetic correlation with ANX, while substantial, is not at unity, and the correlation with PTSD is somewhat lower, supporting the conceptualization of SAD as a partially distinct entity rather than a variant expression of general anxiety. The particularly salient association with sensitivity and hurt feelings, a facet of neuroticism related specifically to interpersonal rejection, is apt given the centrality of fear of negative evaluation to SAD’s phenomenology [2], and this association is less significant in the ANX genetic correlation.

We find that rural residence is associated with substantially increased SAD risk, an association that is robust to adjustment for age, sex, population stratification, and socioeconomic factors. The observed ∼43% increase in SAD odds in the fully adjusted model is contrary to classical urban stress models, which predict a net protective effect of rural environments through reduced social density and environmental stimulation [21,22]. Instead, these findings are more consistent with the social facilitation hypothesis, which states that limited and less varied social exposure in rural settings restricts habituation to evaluative social situations, reinforces avoidance behavior, and thereby sustains and amplifies social anxiety in genetically predisposed individuals [75]. The additional possibility that tighter social surveillance networks in rural communities increase the salience and persistence of negative social evaluation, a core cognitive mechanism in SAD maintenance models, may further explain why there is an elevated genetic risk in this environment. Notably, the minimal attenuation of the effect following SES adjustment suggests that the rural–SAD association cannot merely be attributed to socioeconomic disadvantage alone. The consistency of direction across ancestry groups reinforces that this association is not specific to a single population and is unlikely to be explained purely by differential population stratification.

The heterogeneity analyses stratified by rural and urban residence revealed the locus rs2328715 in the *CASC15* gene as the strongest heterogenous signal. There has also been evidence that the mouse equivalent of this gene is involved in synaptic neuroplasticity, and further studies showing its presence have displayed evidence of learning and memory enhancement in mice [76]. Another heterogenous signal in the rural vs urban analysis, *LMX1B,* is a transcription factor that plays a critical role in the development and maintenance of central serotonergic neurons [77], a system previously linked to SAD [78]. Unique to the rural GWAS, loci mapping to *GABRA4* and *KCTD16*, genes involved in GABAergic neurotransmission, were most notable. Mice studies, particularly, have emphasized the involvement of GABA receptors in fear and anxiety [79–84]. In humans, these genes may fit well within established neurobiological models of anxiety disorders, which emphasize altered inhibitory signaling and exaggerated threat processing [3,87,88]. These genes being unique to the rural GWAS may suggest that the biological pathways of the threat response differ between people living in rural vs urban environments. For instance, granulocyte count, which has been linked to psychological stress and found as significant in a methylwide association study on anxiety, was significant in the rural, but not the urban analyses [91,92]. This is consistent with contemporary models suggesting that environmental context interacts with genetic liability to influence the specific biological mechanisms underlying psychiatric disorders [89,90].

Some limitations should also be acknowledged. Sample sizes for non-EUR ancestry groups and the stratified analyses were substantially smaller, limiting ancestry-specific inference and statistical power to detect population- and environment-specific loci. Phenotype ascertainment relied on ICD-10 codes and survey-derived criteria rather than clinician-administered diagnostic assessment, introducing potential misclassification. PRS analyses were restricted to individuals of EUR ancestry, limiting generalizability and reinforcing the need for multi-ancestry PRS development. Geographic classification using ZIP3-linked RUCA codes may not fully capture within-category environmental heterogeneity, and residual confounding from unmeasured variables, including social capital, community-level mental health stigma, and access to specialist care, which cannot be excluded despite comprehensive socioeconomic covariate adjustment. Future studies incorporating prospective residential history data and functional social exposure measures will be essential to characterize the developmental timing and mechanisms through which geographic context moderates genetic liability for SAD. Rather than treating environmental exposures solely as covariates, approaches that explicitly model GxE interplay may yield deeper insight into the mechanisms through which genetic risk is realized.

## Conclusion

This study demonstrates that SAD may have a partially distinct genetic architecture within the anxiety spectrum, as evidenced by differences in the phenotypic architecture associated with both disorders. Geographic context may also modulate SAD risk in a manner that challenges purely urban-centric models of psychiatric vulnerability. Future research should prioritize adequately powered cross-ancestry GWAS to enable discovery of SAD-specific loci, longitudinal cohort designs to disentangle the developmental timing of environmental exposures and genetic risk expression, and formal GxE interaction analyses to determine whether genetic liability for SAD differs across geographic and socioeconomic contexts. Functional characterization of the identified loci, particularly *NEGR1,* will be essential to translate these associations into mechanistic insight and, ultimately, to inform targeted intervention strategies for at-risk populations.

## Data Availability

All data produced in the present study are available upon reasonable request to the authors.

## Acknowledgements

This work is supported by National Institutes of Health (Grant No. R01MH137212 to RLK, T32HG009495 to KLF). We acknowledge the Penn Medicine BioBank (PMBB) for providing data and thank the patient-participants of Penn Medicine who consented to participate in this research program. We would also like to thank the Penn Medicine BioBank team and Regeneron Genetics Center for providing genetic variant data for analysis. The PMBB is approved under IRB protocol# 813913 and supported by Perelman School of Medicine at University of Pennsylvania, a gift from the Smilow family, and the National Center for Advancing Translational Sciences of the National Institutes of Health under CTSA award number UL1TR001878.

