## Supplementary Material for "Genetic architecture of social anxiety disorder: examining moderation by rural-urban residential environment"

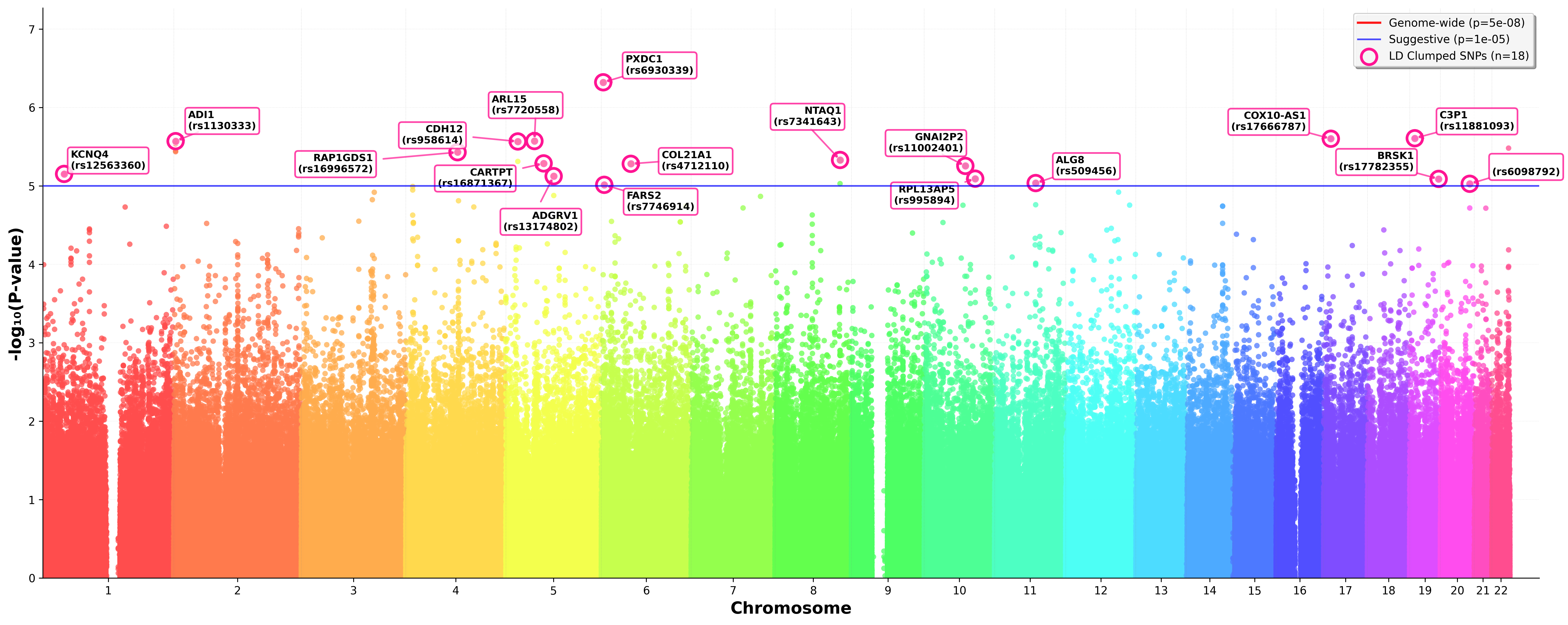
**Supplementary Figure 1.** Manhattan plot showing genome-wide association results for the social anxiety phenotype in the All of Us cohort (ICD10 and/or answering ‘yes’ on three social anxiety survey questions) in African American (AFR) ancestry only. The y-axis represents −log₁₀(p-values), and the x-axis represents chromosomal positions. The solid blue line denotes suggestive significance (p<1×10⁻⁵). Top loci are annotated with nearest gene symbols.
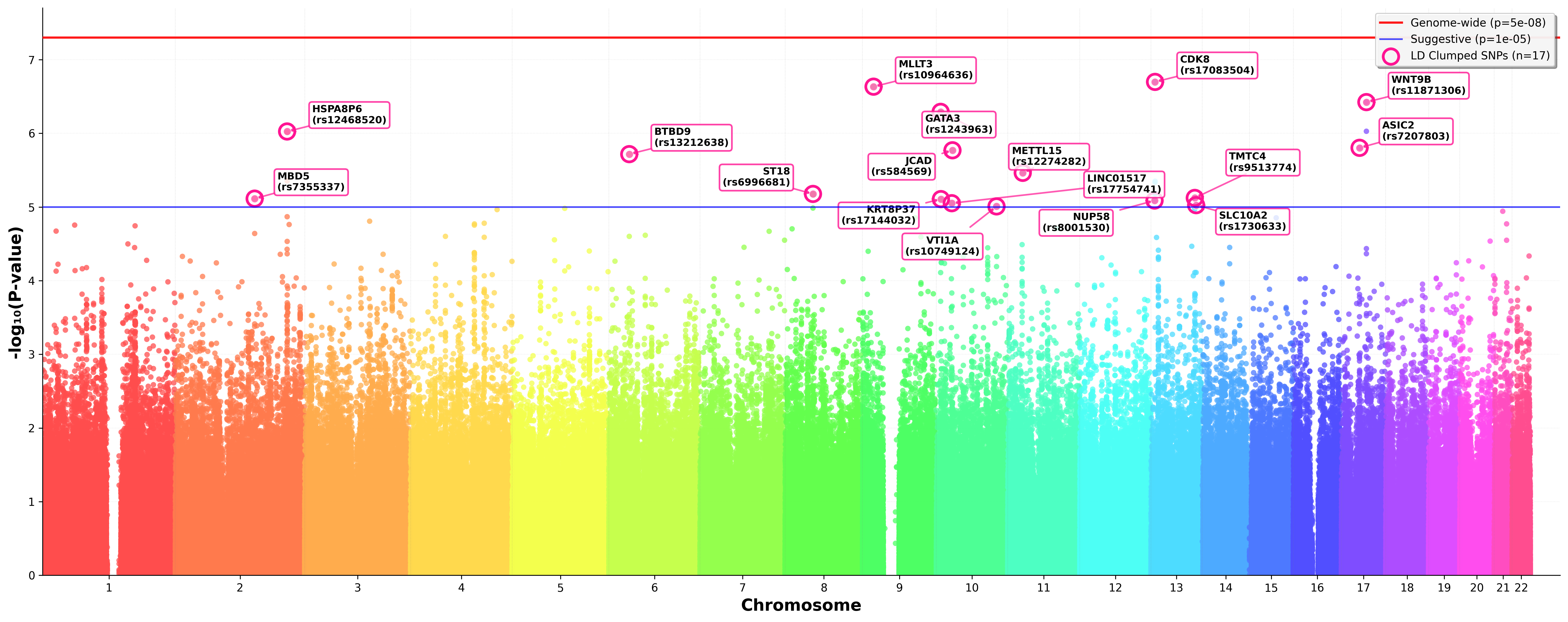
**Supplementary Figure 2.** Manhattan plot showing genome-wide association results for the social anxiety phenotype in the All of Us cohort (ICD10 and/or answering ‘yes’ on three social anxiety survey questions) in Admixed American (AMR) ancestry only. The y-axis represents −log₁₀(p-values), and the x-axis represents chromosomal positions. The solid red line denotes genome-wide significance (p<5×10⁻⁸); the solid blue line denotes suggestive significance (p<1×10⁻⁵). Top loci are annotated with nearest gene symbols.

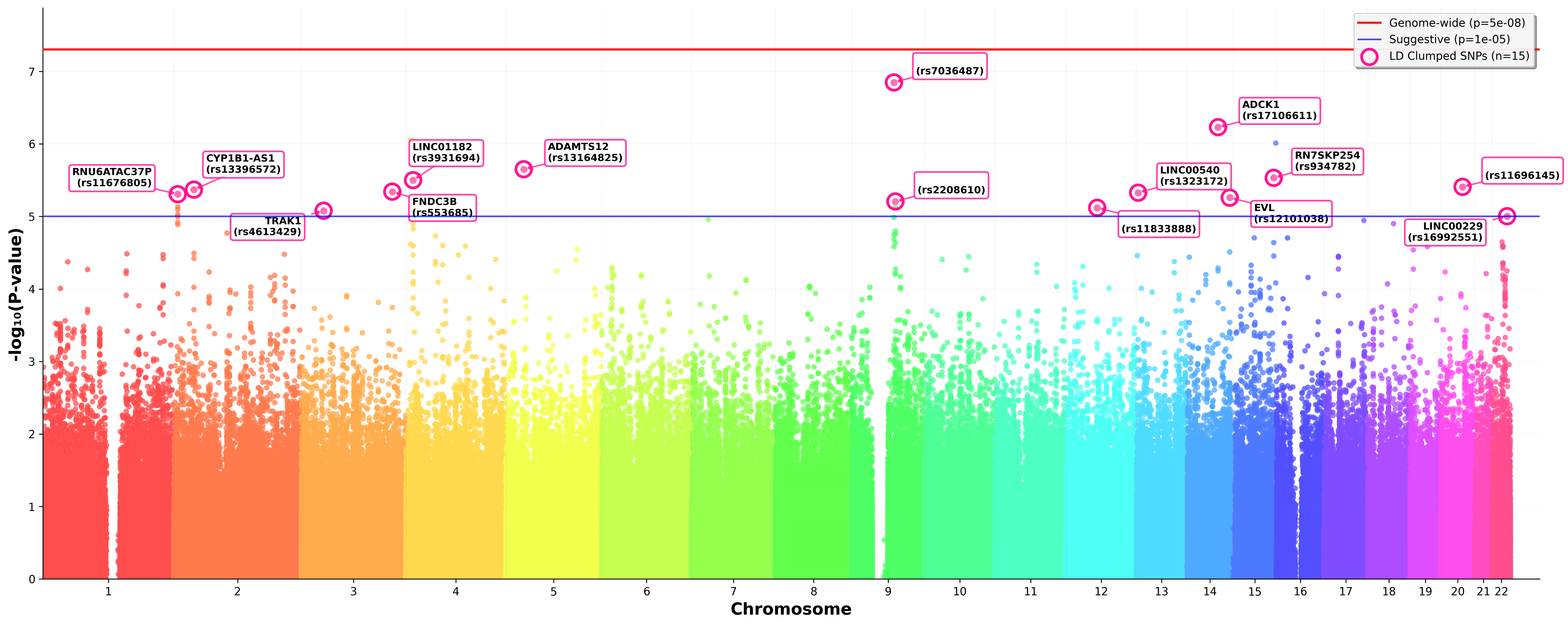
**Supplementary Figure 3.** Manhattan plot showing genome-wide association results for the social anxiety phenotype in the All of Us cohort (ICD10 and/or answering ‘yes’ on three social anxiety survey questions) in East Asian (EAS) ancestry only. The y-axis represents −log₁₀(p-values), and the x-axis represents chromosomal positions. The solid red line denotes genome-wide significance (p<5×10⁻⁸); the solid blue line denotes suggestive significance (p<1×10⁻⁵). Top loci are annotated with nearest gene symbols.

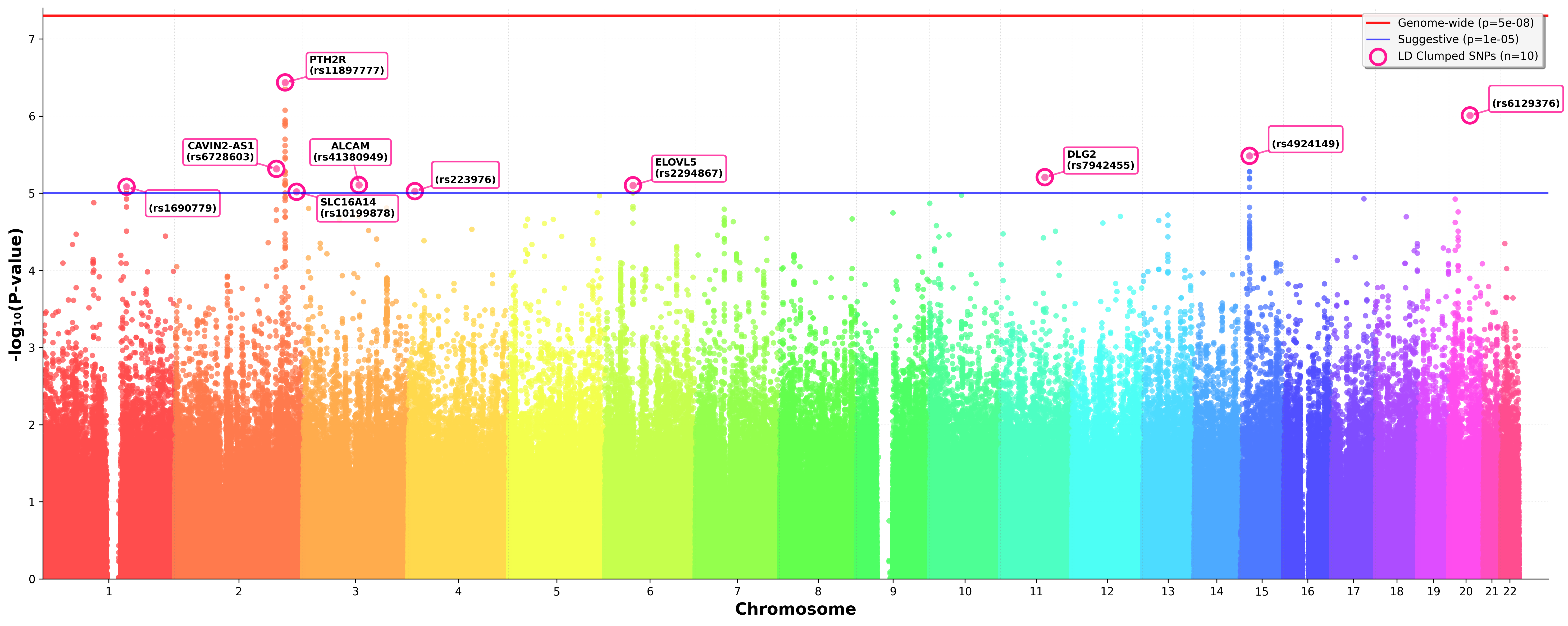
**Supplementary Figure 4.** Manhattan plot showing genome-wide association results for the social anxiety phenotype in the All of Us cohort (ICD10 and/or answering ‘yes’ on three social anxiety survey questions) in European (EUR) ancestry only. The y-axis represents −log₁₀(p-values), and the x-axis represents chromosomal positions. The solid red line denotes genome-wide significance (p<5×10⁻⁸); the solid blue line denotes suggestive significance (p<1×10⁻⁵). Top loci are annotated with nearest gene symbols.

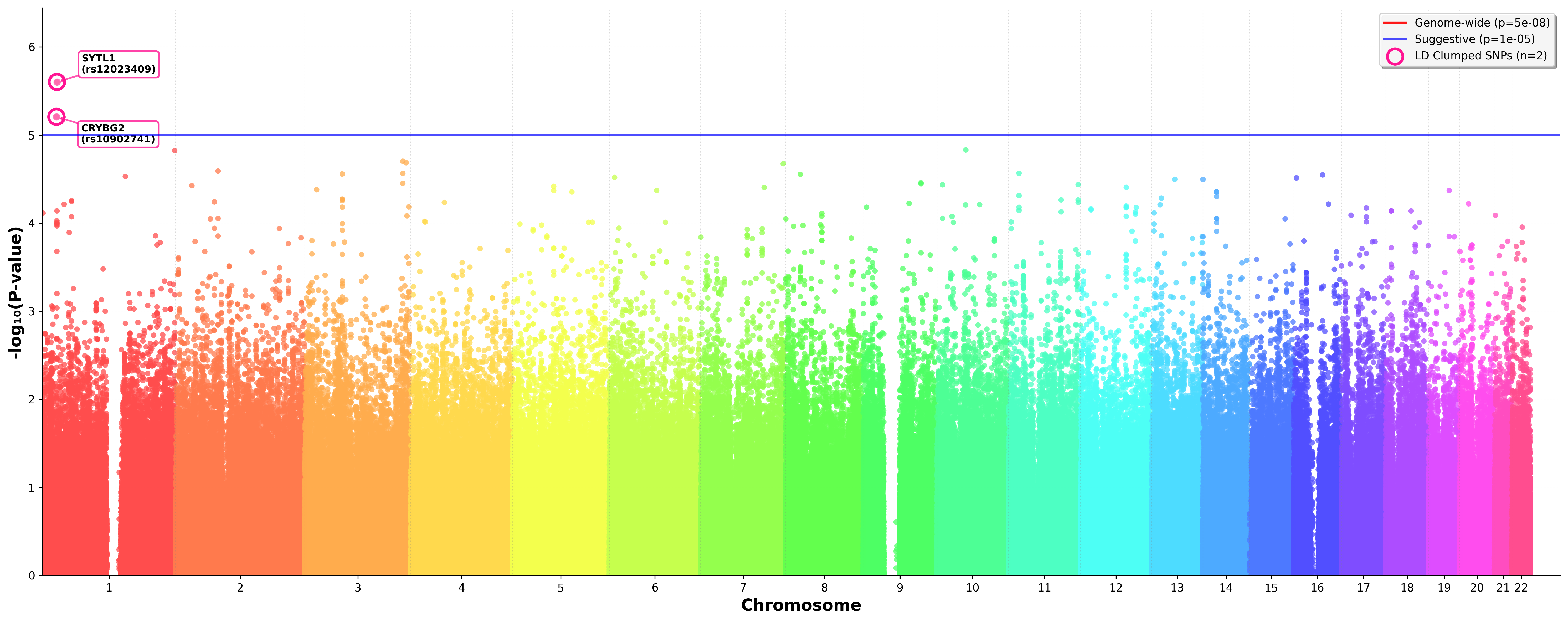
**Supplementary Figure 5.** Manhattan plot showing genome-wide association results for the social anxiety phenotype in the All of Us cohort (ICD10 and/or answering ‘yes’ on three social anxiety survey questions) in South Asian (SAS) ancestry only. The y-axis represents −log₁₀(p-values), and the x-axis represents chromosomal positions. The solid blue line denotes suggestive significance (p<1×10⁻⁵). Top loci are annotated with nearest gene symbols.

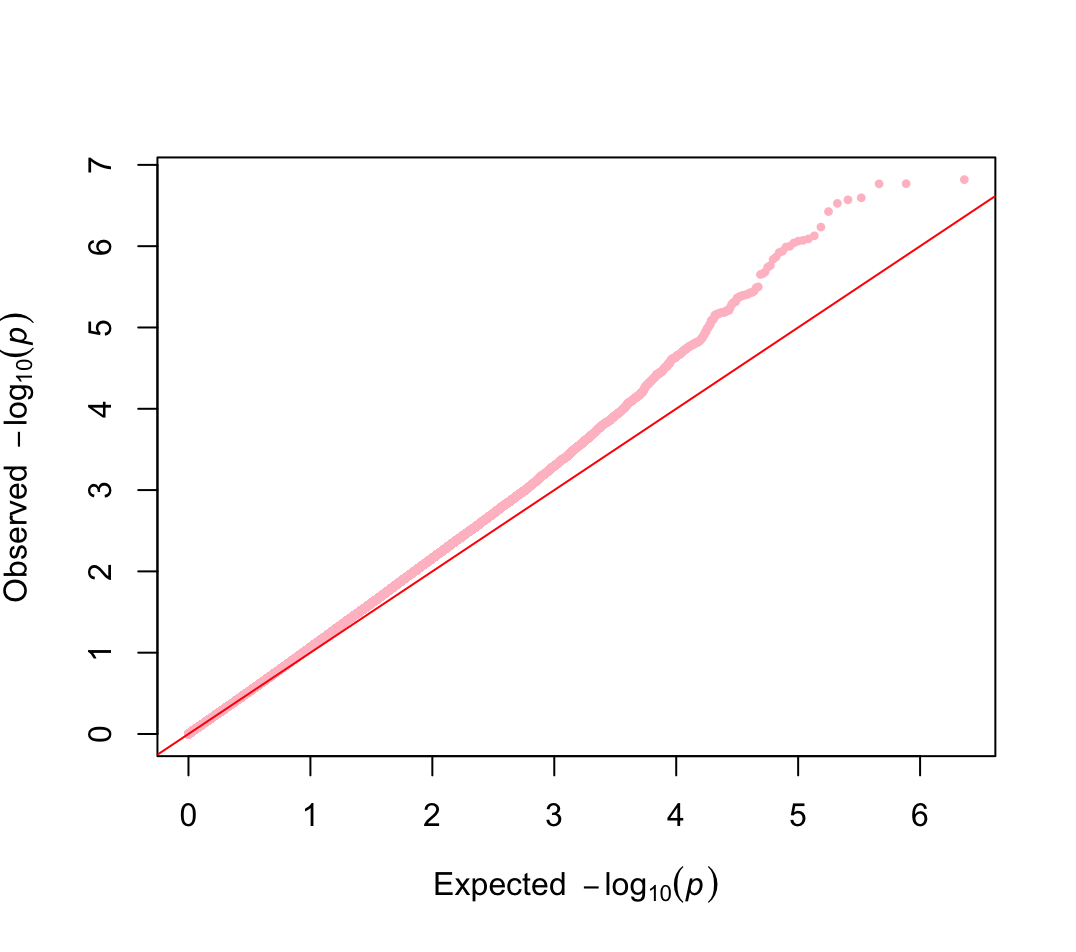
**Supplementary Figure 6.** A quantile-quantile (QQ) plot of the meta-analyzed cross-ancestry AoU GWAS p-values. The calculated lambda (λ) value was 1.08.

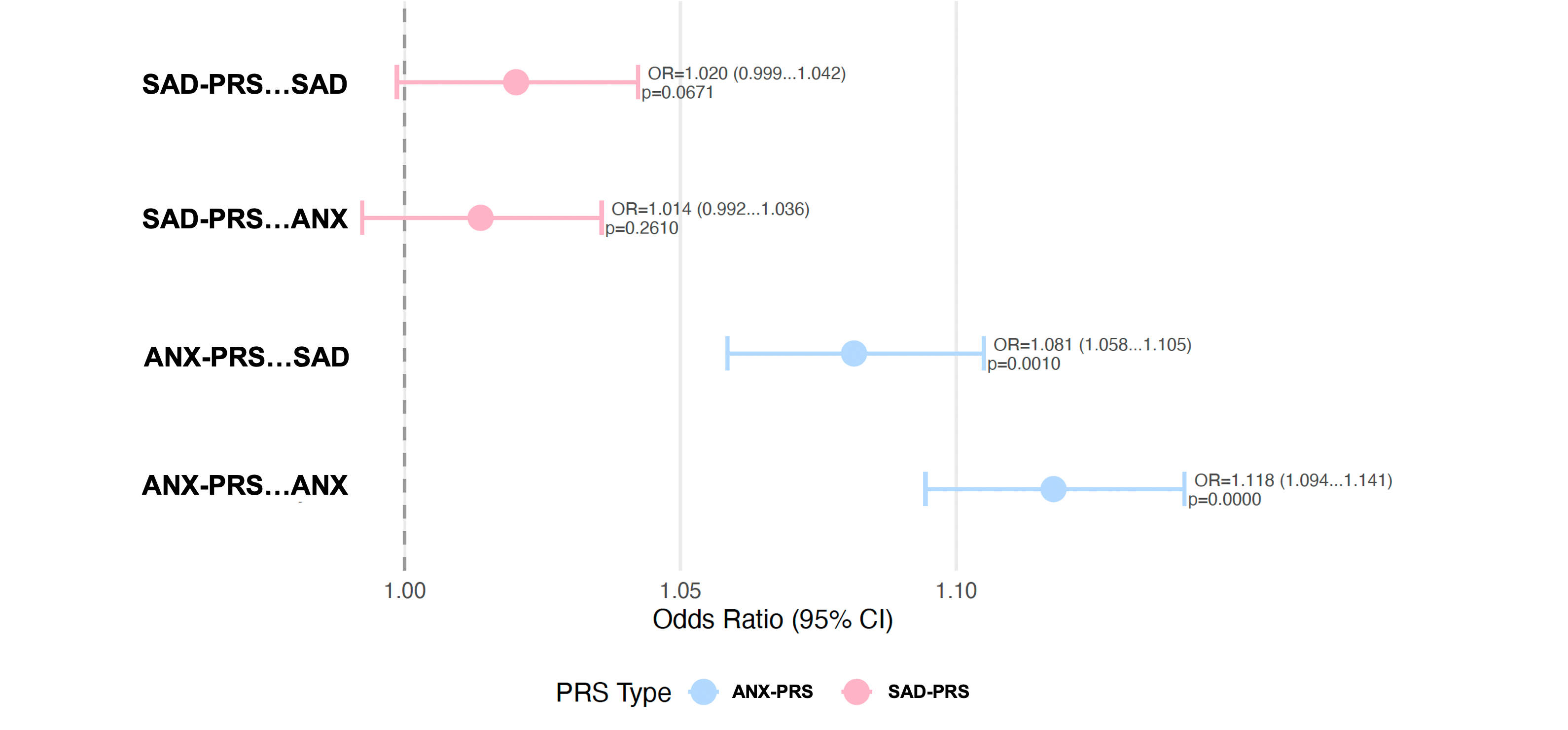

**Supplementary Figure 7.** Forest plot showing associations between polygenic risk scores (PRSs) and clinical phenotypes of major anxiety disorders (ANX) and social anxiety disorder (SAD). Points represent odds ratios (ORs), and horizontal bars indicate 95% confidence intervals (CIs). The dashed vertical line denotes the null association (OR = 1.0). Blue markers correspond to GA-PRS associations and pink markers correspond to SAD-PRS associations. ANX-PRS demonstrated significant positive associations with both ANX phenotypes (OR = 1.081, 95% CI: 1.058-1.105, p = 0.0010; OR = 1.118, 95% CI: 1.094-1.141, p < 0.0001), whereas associations involving SAD-PRS were weaker and did not reach statistical significance (p = 0.0671 and p = 0.2610).

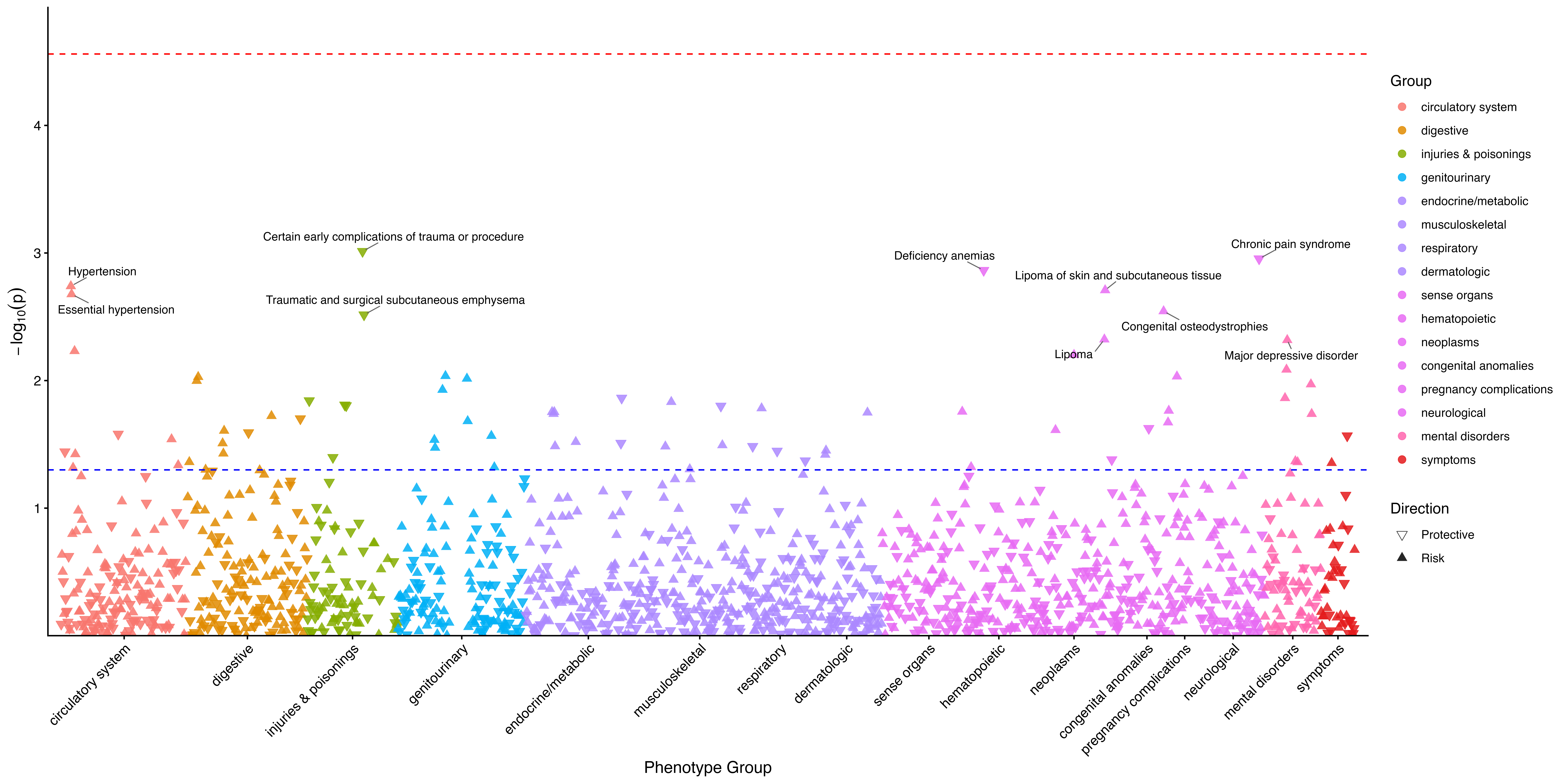

**Supplementary Figure 8.** Phenome-wide association study (PheWAS) results assessing associations between polygenic risk and clinical phenotypes across disease categories in females only. Each point represents an individual phenotype in the Phecode 1.2 disease system (with a minimum of two phecodes and exclusion map true) and colored by phenotype category. Upward-pointing triangles indicate risk associations, whereas downward-pointing triangles indicate protective associations.

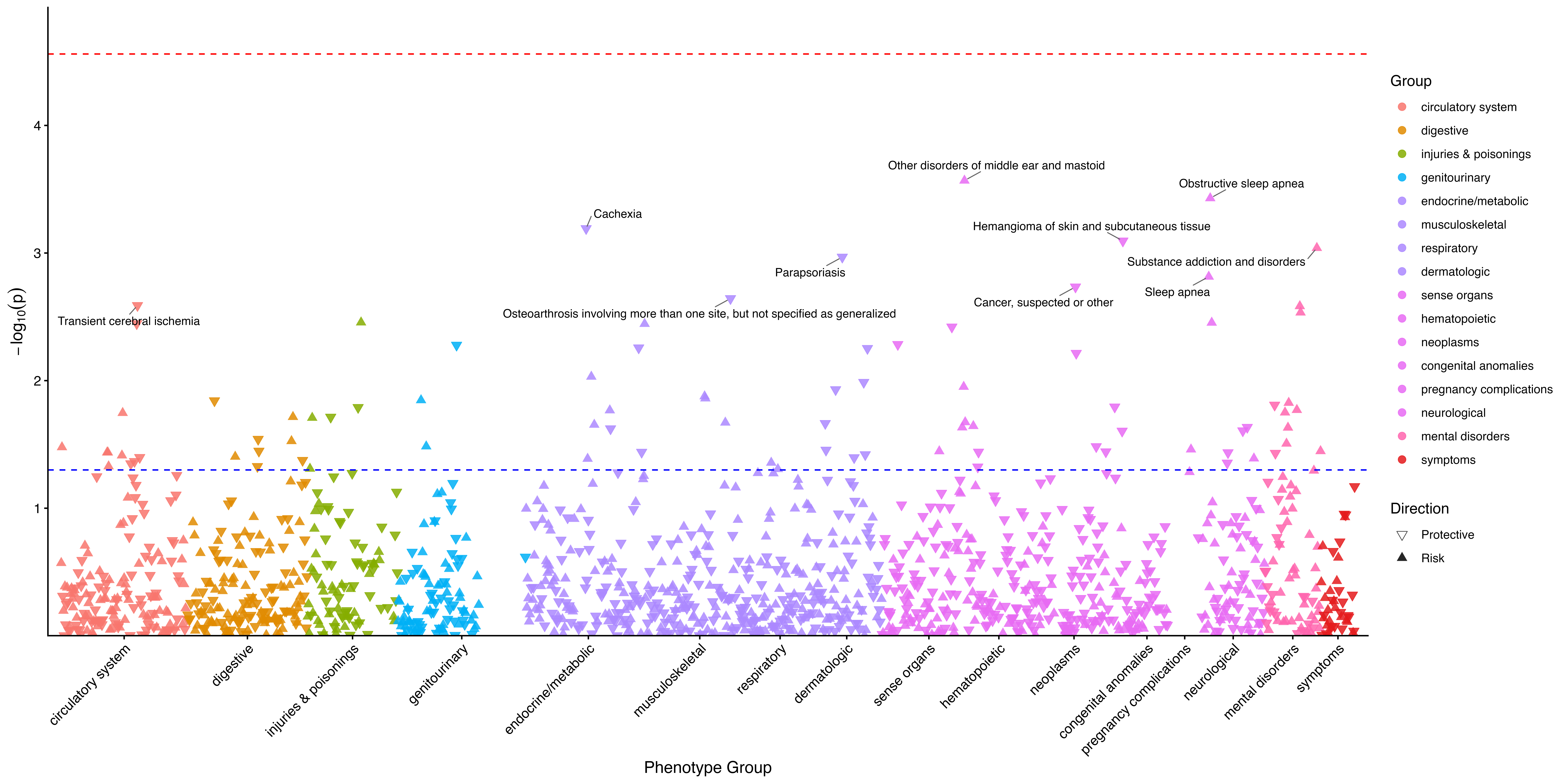
**Supplementary Figure 9.** Phenome-wide association study (PheWAS) results assessing associations between polygenic risk and clinical phenotypes across disease categories in males only. Each point represents an individual phenotype in the Phecode 1.2 disease system (with a minimum of two phecodes and exclusion map true) and colored by phenotype category. Upward-pointing triangles indicate risk associations, whereas downward-pointing triangles indicate protective associations.

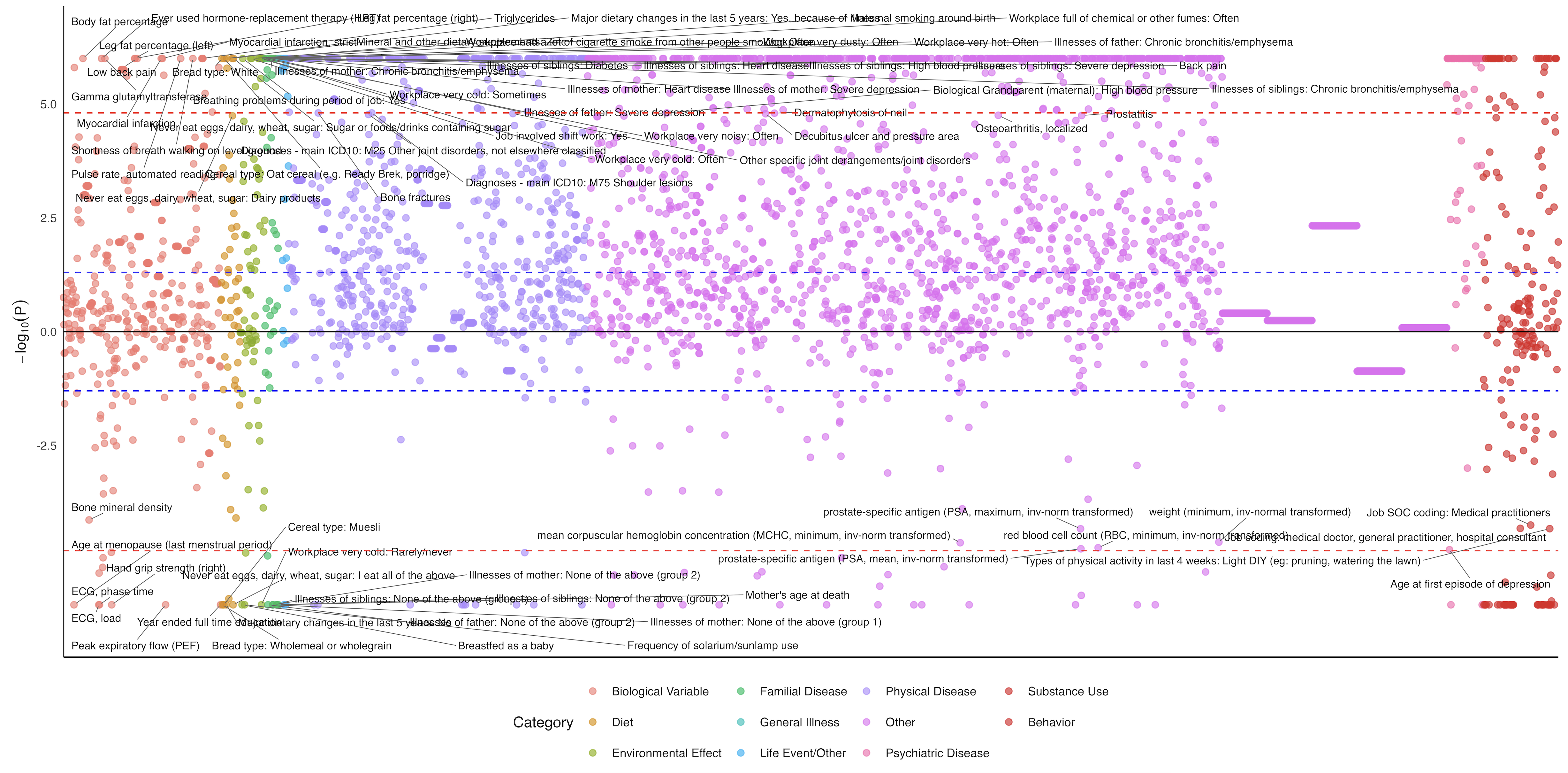

**Supplementary Figure 10**. Genetic correlation analysis on different traits compared to major anxiety disorders (ANX), generated by the Complex Traits Genetics Virtual Lab in European ancestry. The y-axis represents the log-transformed false discovery rate–corrected p values across the traits included in the analysis, with the dashed line representing the significance threshold and yellow circles representing those that meet the Bonferroni significance.

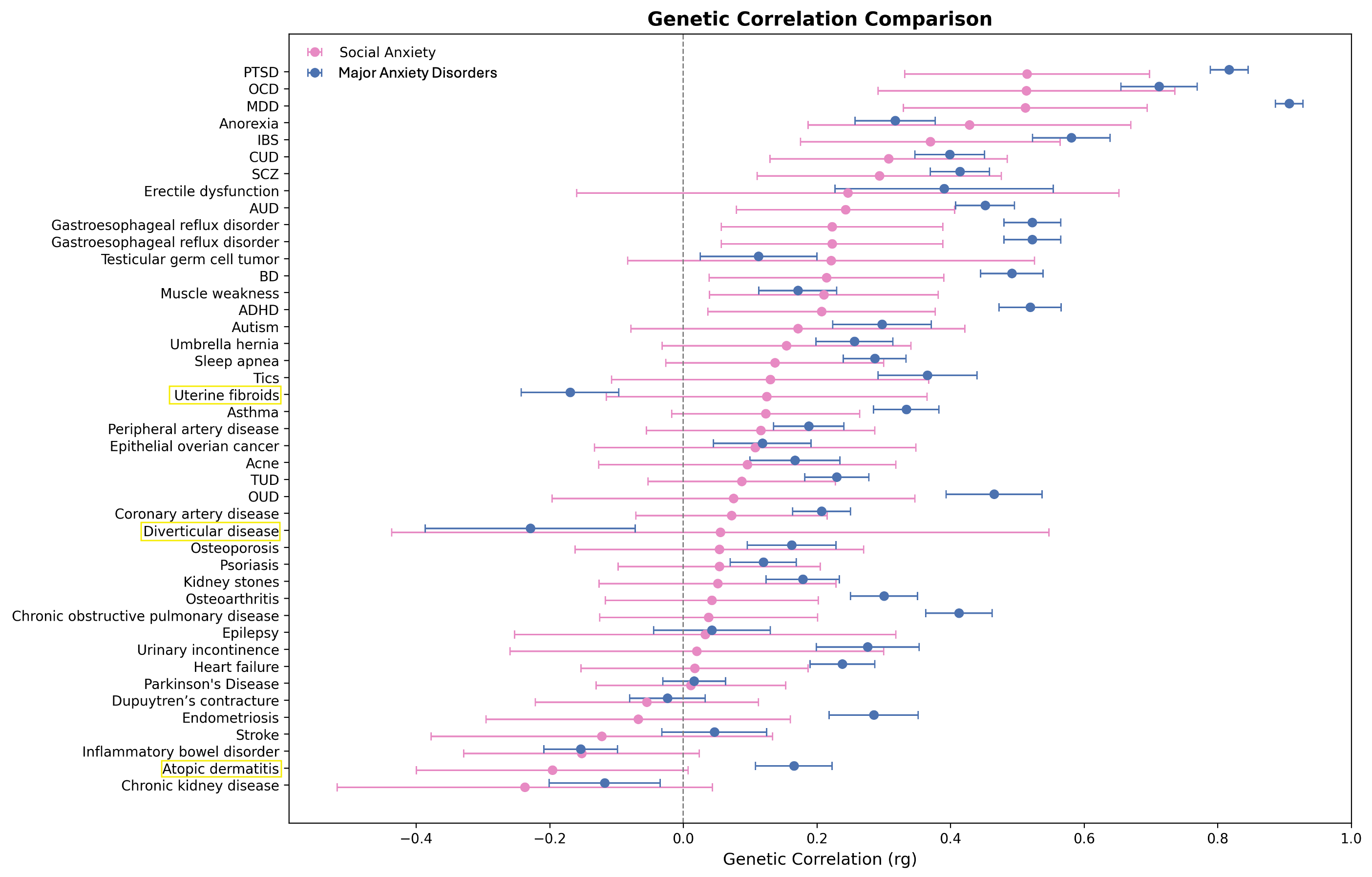
**Supplementary Figure 11**. Forest plot comparing the genetic correlations of the most highly correlated disorders shared between SAD (pink) and major anxiety disorders (ANX) (blue). Highlighted in yellow boxes are those where the direction between SAD and ANX differs.

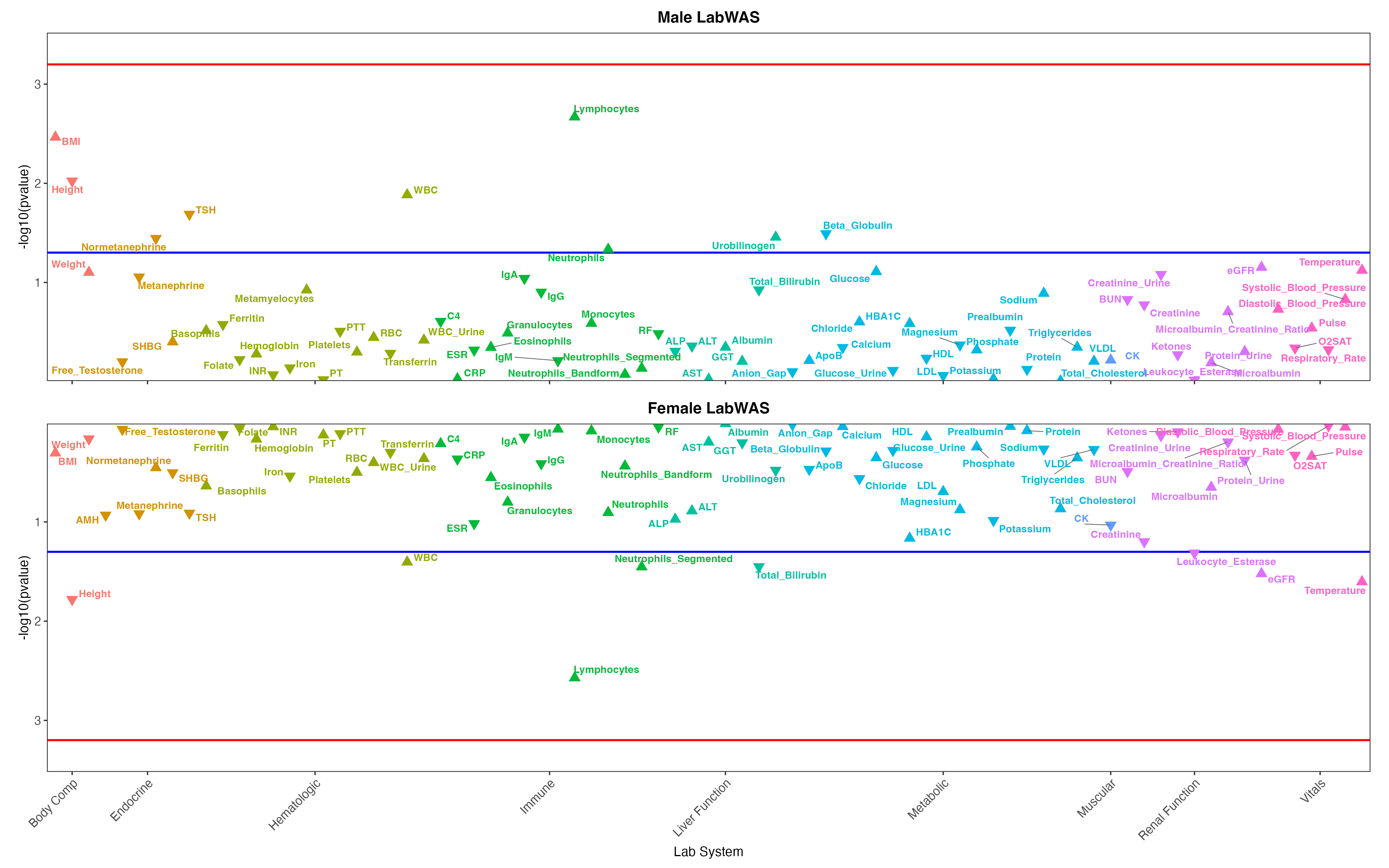
**Supplementary Figure 12.** Hudson plot showing laboratory-wide association study (LabWAS) examining associations between social anxiety disorder (SAD) polygenic risk scores and laboratory traits among European males vs European females. Each point represents a laboratory measurement, grouped and color-coded according to physiological system. Each laboratory measure is represented by a directional triangle, where ▲ indicates a positive effect estimate (higher laboratory values associated with greater disorder risk) and ▼ indicates a negative effect estimate (lower laboratory values associated with greater disorder risk). The blue horizontal line indicates the nominal significance threshold (p = 0.05), and the red horizontal line indicates the multiple-testing correction threshold (P < 0.05 / 79 tests). WBC indicates white blood cell count; eGFR estimated Glomerular Filtration Rate; HBA1C Hemoglobin A1c; TSH Thyroid-Stimulating Hormone.

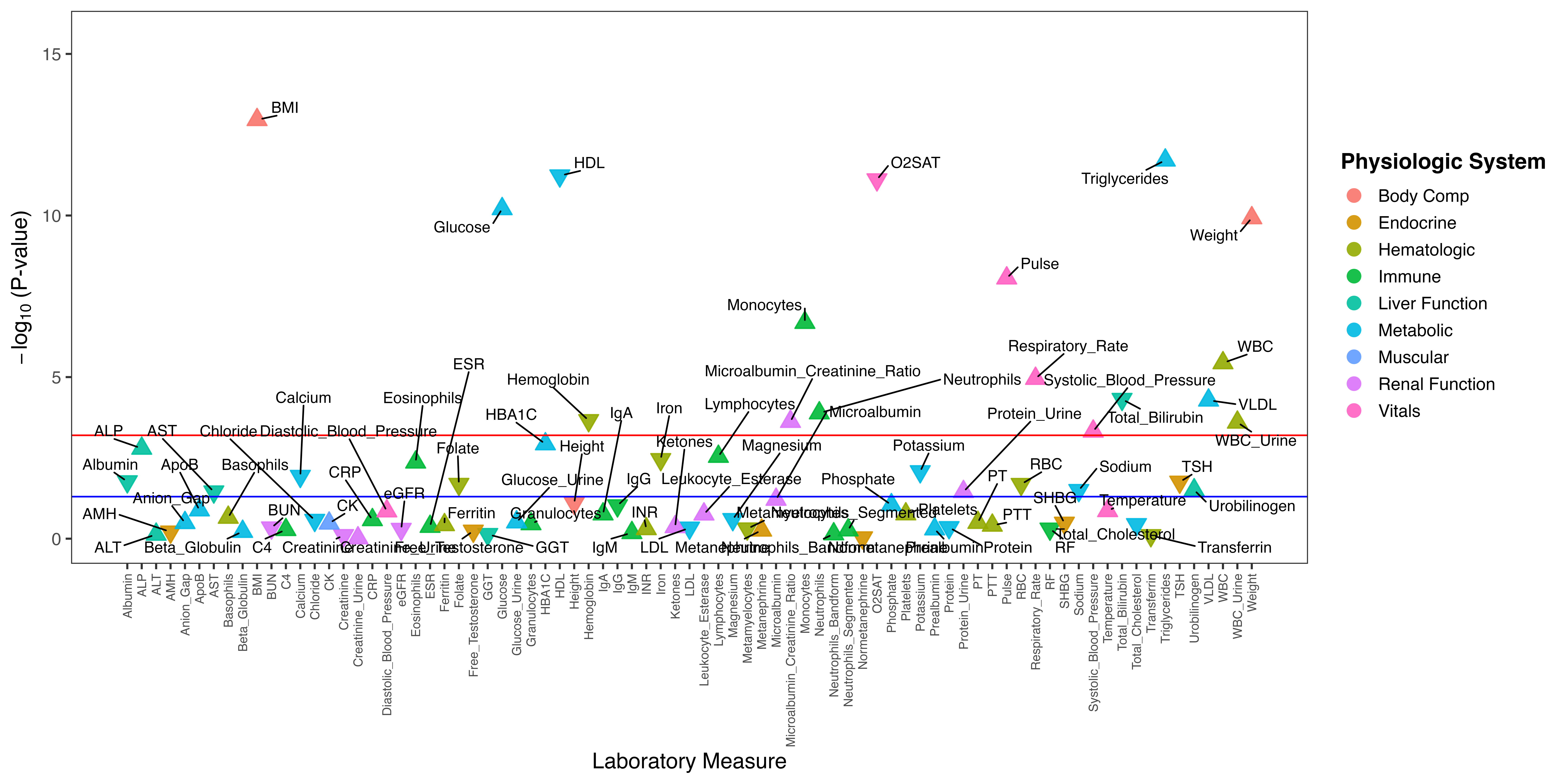

**Supplementary Figure 13**. Laboratory-wide association study (LabWAS) examining associations between major anxiety disorders (ANX) polygenic risk scores and laboratory traits among Europeans. Each point represents a laboratory measurement, grouped and color-coded according to physiological system. Each laboratory measure is represented by a directional triangle, where ▲ indicates a positive effect estimate (higher laboratory values associated with greater disorder risk) and ▼ indicates a negative effect estimate (lower laboratory values associated with greater disorder risk). The blue horizontal line indicates the nominal significance threshold (p = 0.05), and the red horizontal line indicates the multiple-testing correction threshold (P < 0.05 / 79 tests). WBC indicates white blood cell count; eGFR estimated Glomerular Filtration Rate; HBA1C Hemoglobin A1c; TSH Thyroid-Stimulating Hormone.

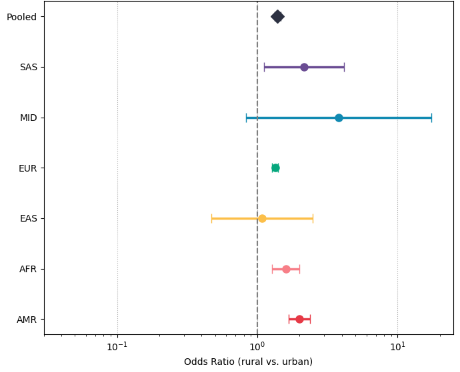

**Supplementary Figure 14**. Forest plot showing the odds of social anxiety per ancestry in rural vs urban (pooled = black, SAS [n=5368] = purple, MID [n=1467] = blue, EUR [n=222,500] = green, EAS [n=9707] yellow, AFR [n=74,576] = orange, AMR [n=70,626] = red), with a higher odds in a rural environment being >1 and higher odds in an urban environment being <1.

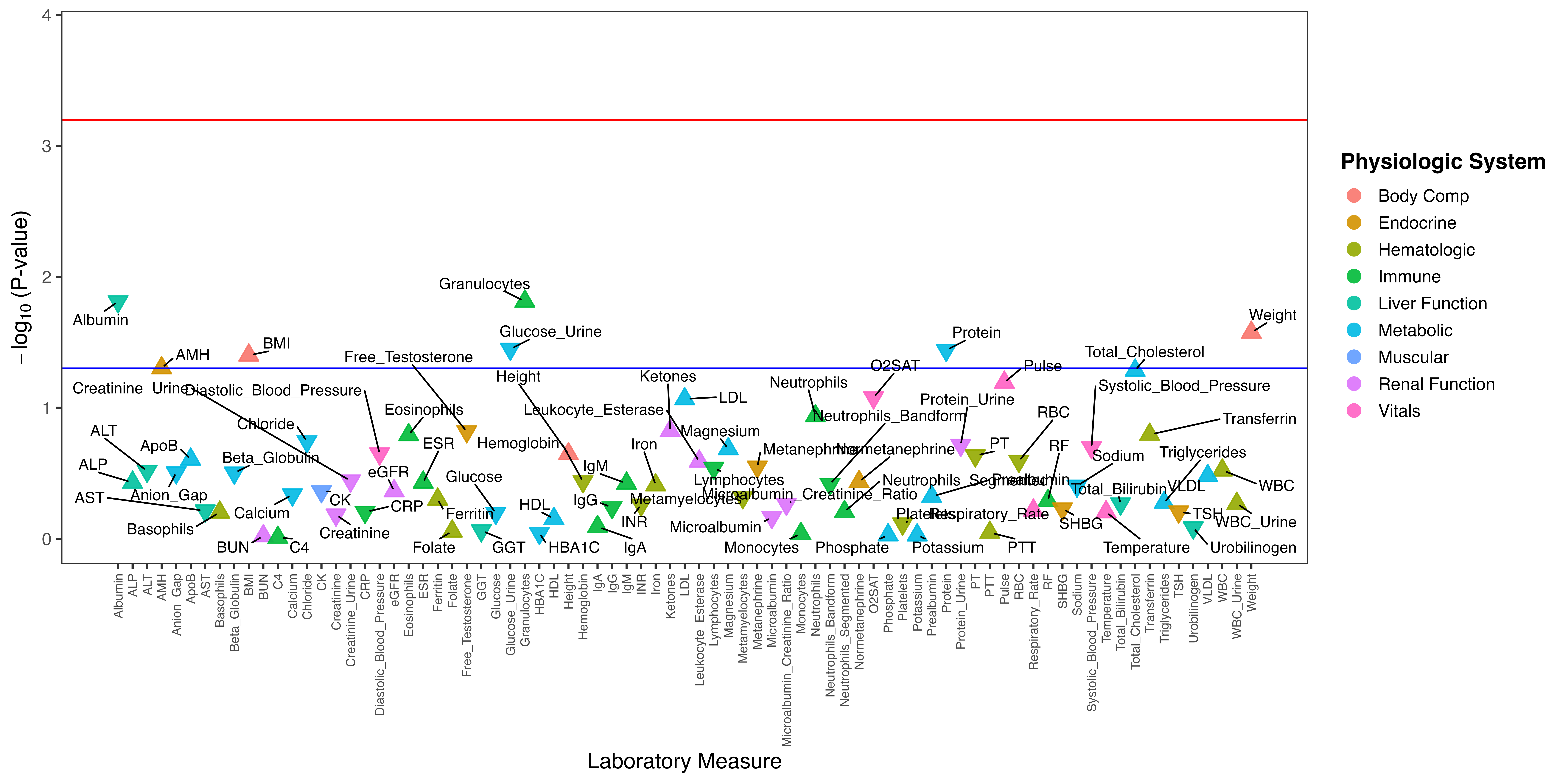

**Supplementary Figure 15**. Laboratory-wide association study (LabWAS) examining associations between all SAD polygenic risk scores and laboratory traits among Europeans in rural residence. Each point represents a laboratory measurement, grouped and color-coded according to physiological system. Each laboratory measure is represented by a directional triangle, where ▲ indicates a positive effect estimate (higher laboratory values associated with greater disorder risk) and ▼ indicates a negative effect estimate (lower laboratory values associated with greater disorder risk). The blue horizontal line indicates the nominal significance threshold (p = 0.05), and the red horizontal line indicates the multiple-testing correction threshold (P < 0.05 / 79 tests). WBC indicates white blood cell count; eGFR estimated Glomerular Filtration Rate; HBA1C Hemoglobin A1c; TSH Thyroid-Stimulating Hormone.

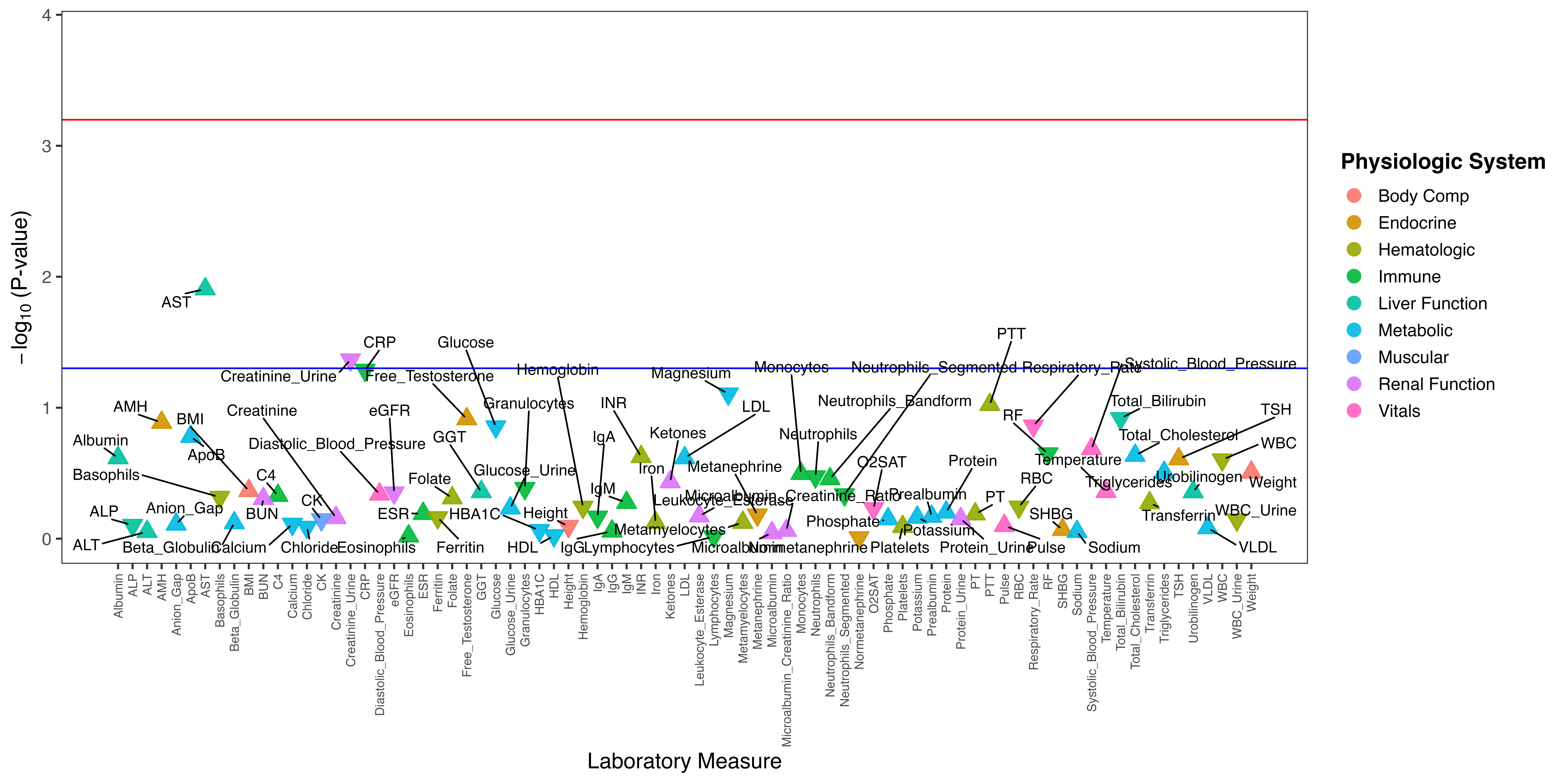

**Supplementary Figure 16**. Laboratory-wide association study (LabWAS) examining associations between all SAD polygenic risk scores and laboratory traits among Europeans in urban residence. Each point represents a laboratory measurement, grouped and color-coded according to physiological system. Each laboratory measure is represented by a directional triangle, where ▲ indicates a positive effect estimate (higher laboratory values associated with greater disorder risk) and ▼ indicates a negative effect estimate (lower laboratory values associated with greater disorder risk). The blue horizontal line indicates the nominal significance threshold (p = 0.05), and the red horizontal line indicates the multiple-testing correction threshold (P < 0.05 / 79 tests). WBC indicates white blood cell count; eGFR estimated Glomerular Filtration Rate; HBA1C Hemoglobin A1c; TSH Thyroid-Stimulating Hormone.

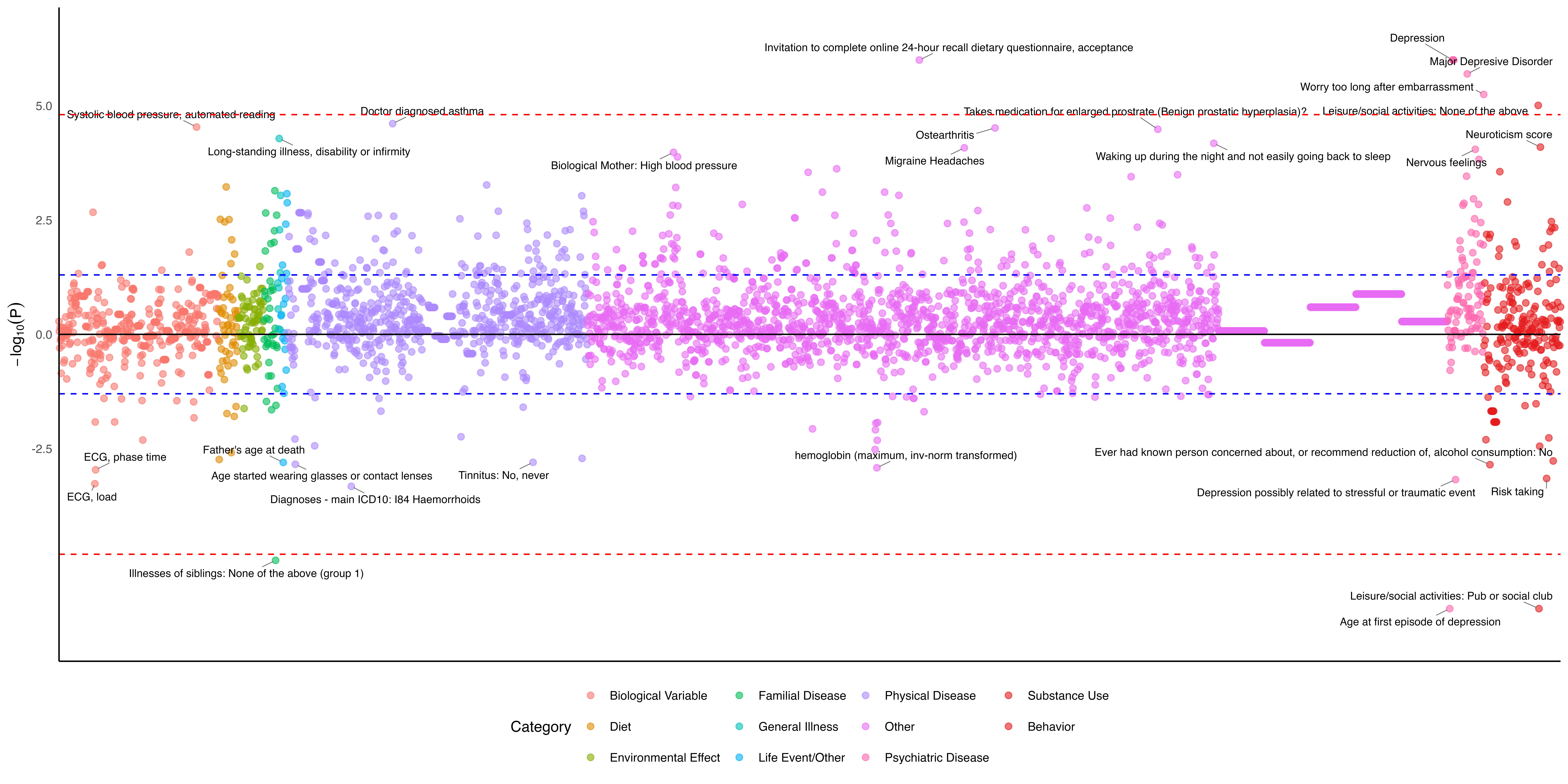

**Supplementary Figure 17**. Genetic correlation analysis on different traits compared to SAD in urban residence, generated by the Complex Traits Genetics Virtual Lab in European ancestry. The y-axis represents the log-transformed false discovery rate–corrected p values across the traits included in the analysis, with the dashed line representing the significance threshold and yellow circles representing those that meet the Bonferroni significance.
